# Genetic Spectrum of Autism Spectrum Disorder Associated Variants in the Indian Population: Insights from 1,029 IndiGenomes

**DOI:** 10.1101/2025.08.14.25333641

**Authors:** Mukesh Kumar, Treesa Issen, Mohammed Swalih, Srishti Sharma, Sheffali Gulati, Sridhar Sivasubbu, Vinod Scaria, BK Binukumar

## Abstract

**Background:** Autism spectrum disorder (ASD) is a lifelong neurodevelopmental condition characterised by social and behavioural challenges. It is highly heritable, with estimates ranging from 40-90% and a liability-scale heritability of 0.924, underscoring the role of genetic variants in its pathogenesis. Limited studies from India hinder an accurate assessment of the country’s true ASD genetic spectrum.

**Aim:** To investigate the genetic spectrum in an Indian control dataset, providing baseline variant frequency data to facilitate interpretation of disease-associated variants in the Indian population.

**Methods:** We compiled a strongly ASD-linked gene panel from the SFARI and ClinGen databases, followed by the extraction of gene-associated variants from the IndiGenomes dataset. A stepwise analysis pipeline was implemented, including variant annotation, filtering for minor allele frequency (MAF < 5%), prediction of deleteriousness using REVEL (>0.75), and loss-of-function prediction with LoFTEE (high confidence). Variants were then classified according to ACMG-AMP guidelines.

**Results:** We identified 1,531,327 variants from 517 ASD panel genes (MAF < 5%) spanning ∼88 Mb of genomic regions. ACMG-AMP interpretation was performed for prioritised 298 missense variants (REVEL > 0.75) and 537 LoF variants (LoFTEE; high confidence). Missense and LoF variants contributed 37% and 63% of the filtered data, respectively. Fifty-five variants from 28 genes were classified as pathogenic/likely pathogenic, while 10 variants in 6 genes showed significantly higher MAF (p < 0.05; Fisher’s exact test). Carrier numbers were estimated for autosomal recessive inheritance models.

**Conclusion:** This study provides a comprehensive spectrum of ASD-related genetic variants in the Indian population, offering valuable insights for ASD diagnosis, genetic interpretation, and understanding of underlying molecular mechanisms.

## Introduction

The term "autism" originally described self-withdrawal in schizophrenia but was redefined in the 1940s by Dr. Leo Kanner and Dr. Hans Asperger to describe children with social, communication, and repetitive behavioural challenges [1]. Their work, influenced by Georg Frankl’s idea of disrupted ‘affective language’ and ‘word language’, helped define autism as a neurobiological spectrum, now recognised as Autism Spectrum Disorder (ASD) under Diagnostic and Statistical Manual of Mental Disorders (DSM-5; a reference book on mental health and brain-related conditions and disorders) [2]. ASD includes neurodevelopmental conditions affecting social interaction, communication, and behaviour, encompassing autism, Asperger syndrome, PDD-NOS, and related genetic conditions [3].

Global ASD prevalence is 0.77%, with higher rates (1.14%) observed in males [4]. The four studies, including rural (4,750 children) and urban populations totalling 125,849 children, highlight urban-focused ASD prevalence research. The pooled ASD prevalence was 0.11% (95% CI: 0.01–0.20) in rural children (1–18 years old) and 0.09% (95% CI: 0.02–0.16) in urban children (0–15 years old) [5–9]. Limited ASD studies in India hinder the understanding of the true national burden [5].

ASD is highly heritable, with a liability-scale heritability of 0.924, with twin and family studies estimating heritability between 40% and 90%[10,11]. Genetic causes, including de novo mutations and rare variants (MAF ≤ 0.1%) and common variants (small effect), explain 20–25% of ASD cases [10,12–15]. In a cohort of 199 individuals (144 ASD cases and 55 siblings), 243 rare de novo variants (DNVs) were identified. Rare de novo variants (DNVs) were observed in 71.5% of ASD cases and 63% of unaffected siblings, with a comparable mean DNV rate per child (1.24 in ASD vs. 1.18 in siblings), aligning with previous studies [15]. ASD-associated genes are catalogued in well-known databases, including SFARI [16] and ClinGen [17], though a standardised and stringent, clinically validated ASD gene panel remains lacking.

Despite research identifying many ASD-associated genes, the pathogenic variant spectrum remains unclear, especially in underrepresented populations like India. To address this, we analysed whole-genome sequencing data from 1,029 self-reported healthy individuals in the IndiGenomes project [18]. This study developed a stringent ASD gene panel using the SFARI and ClinGen databases to explore the distribution of rare variants in the underrepresented Indian population.

## METHODS

### IndiGenome population dataset

The IndiGen project involved whole-genome sequencing of 1,029 self-declared healthy individuals from diverse geographical regions across India, representing the country’s broad ancestral diversity. The cohort comprised 495 males (mean age: 41.35 years) and 534 females (mean age: 32.96 years). All participants underwent health screening to exclude major known diseases and provided informed consent per the Institutional Human Ethics Committee (IHEC) guidelines of the CSIR-Institute of Genomics and Integrative Biology. Variant data and allele frequencies from this cohort have been previously published in the IndiGenomes database[18].

### Collation and Prioritisation of Autism Spectrum Disorder Genes

A curated list of ASD-associated genes was compiled from two well-established resources: the Simons Foundation Autism Research Initiative (SFARI) Gene database and the ClinGen Intellectual Disability and Autism Gene Curation Expert Panel. A total of 1333 genes from the SFARI and the ClinGen database. To generate a stringent ASD gene list, we prioritised genes with an EAGLE (Evaluation of Autism Gene Link Evidence; quantitative metric for ASD genes) score, greater than 12, as well as those classified as Syndromic (S; genes cause syndromes often featuring ASD) or High Confidence 1 (HC=1; genes have strong replicated ASD evidence) in the SFARI database, and genes marked as Definitive (DE; genes have long-established, proven disease links) or Strong (convincing but shorter-term supporting evidence) in ClinGen. Genes from ClinGen were excluded if they had an EAGLE score below 12 and lacked both Syndromic and HC1 classification, and a final, non-redundant ASD gene list was compiled (Figure 1).

**Figure 1:**
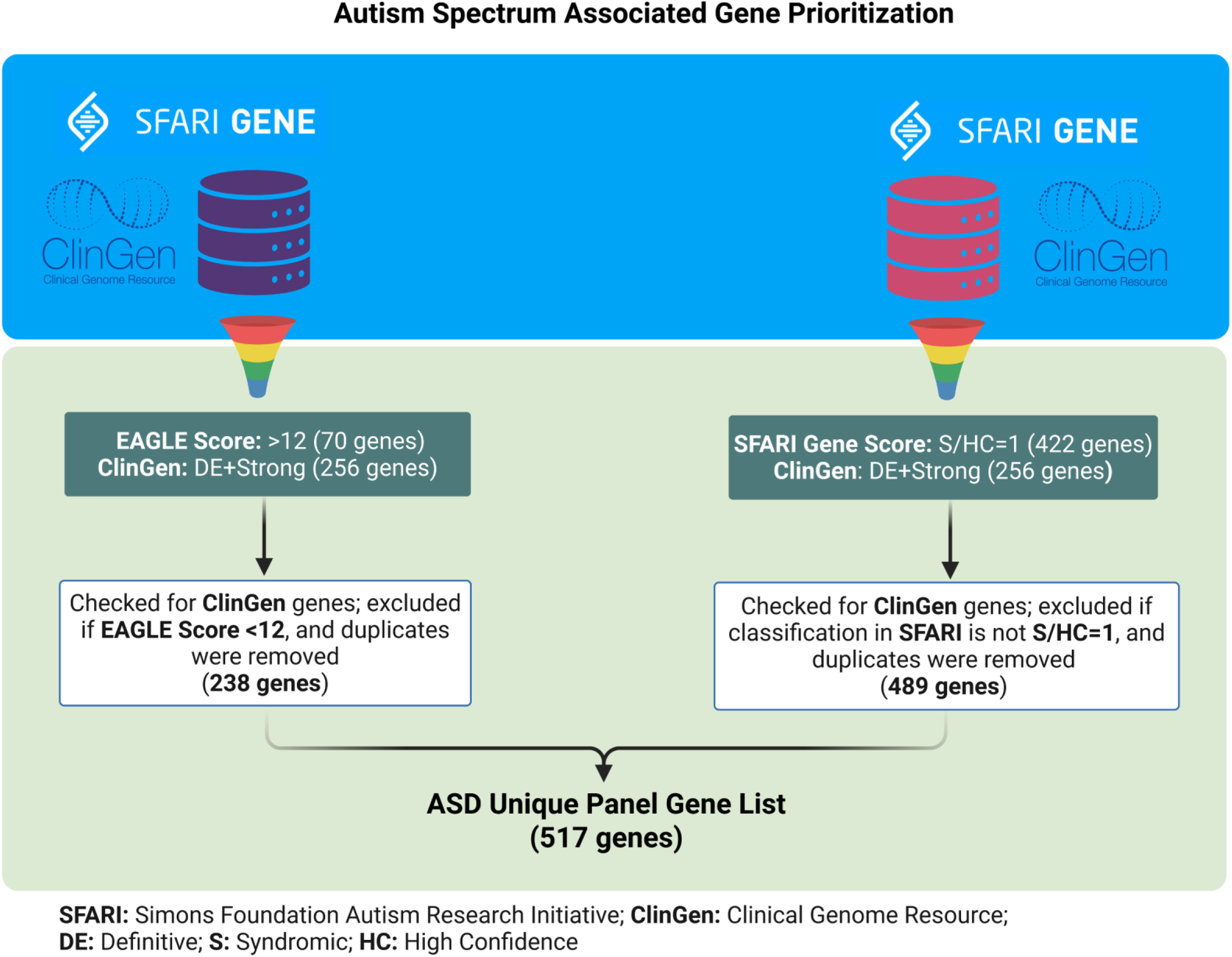
Workflow outlining the gene prioritisation strategy using curated gene lists from the SFARI and ClinGen databases. High-confidence ASD-associated genes were selected through stringent filtering criteria to ensure relevance and specificity.

### Preparation of BED File for Targeted Genomic Analysis of ASD-Associated Genes

The targeted BED file was prepared using Matched Annotation from NCBI and EMBL-EBI (MANE) transcripts, covering the entire gene regions of the prioritised ASD gene panel. BED file generation was performed using a Linux-based command-line interface (CLI), with hg38 refGene annotations downloaded from the NCBI FTP platform (release 1.4). BCFtools was used to extract targeted variants from the IndiGenomes normalised VCF, representing whole-genome data of 1,029 self-reported healthy individuals, using the prepared BED file as the target region input.

### Variant Annotation

Variant annotation was conducted using a customised and updated ANNOVAR pipeline via a Linux-based command-line interface. Gene-based annotations utilised the RefSeq refGene database. Known variants were annotated using avsnp151 (dbSNP build 151). Functional predictions were derived from dbNSFP v4.7a, which integrates multiple in silico tools. Clinical significance was assessed using ClinVar (April 2025 release). Population allele frequencies were annotated using IndiGenomes1029 (Indian dataset), 1000 Genomes (August 2015), gnomAD v4.1 exome/genome (2023), and the GME Variome Project. Splicing effects were assessed using dbscSNV v1.1, while regulatory intronic variants were annotated using regSNP-intron. REVEL scores were used to prioritise potentially damaging missense variants.

Variants were prioritised through a structured, multi-step filtering process. Initially, variants with a minor allele frequency (MAF) greater than 5% in the 1000 Genomes Project, gnomAD exome, and gnomAD genome datasets were excluded to remove common polymorphisms. Synonymous variants and those with unknown consequences were filtered out to focus on potentially functional changes.

### Prioritisation of Missense ASD Gene Panel Variants

Following variant annotation using ANNOVAR, missense variants were extracted using variant class annotation filters from ExonicFunc_refGene. Subsequently, predicted deleterious missense (nonsynonymous) variants were prioritised using the REVEL in-silico predictor, which integrates scores from multiple tools, including MutPred, FATHMM v2.3, VEST 3.0, PolyPhen-2, SIFT, PROVEAN, MutationAssessor, MutationTaster, LRT, GERP++, SiPhy, phyloP, and phastCons[19]. REVEL is a meta-predictor trained on new diseases and harmless variants for better accuracy. Missense variants with a REVEL score greater than 0.75 were retained for further analysis.

### LoF (Loss-of-Function) Variant Annotation and Prioritisation

Loss-of-function (LoF) variants were annotated and assessed for high-confidence (HC) classification using the Variant Effect Predictor (VEP) integrated with the LoFTEE plugin[20]. Annotation was performed by focusing specifically on MANE (Matched Annotation from NCBI and EMBL-EBI) transcripts to ensure consistency and accuracy in transcript selection. High-confidence LoF variants were prioritised for downstream analysis based on LoFTEE’s stringent filtering criteria.

### Classification of Prioritised Variants

Prioritised genetic variants from the IndiGenome control dataset were processed for the classification recommended by ACMG-AMP (American College of Medical Genomics & Genetics - Association of Molecular Pathology) standard guidelines and previously published work [21,22]. Variants were categorised: pathogenic, likely pathogenic, benign, likely benign, and variants of uncertain significance (VUS). In the first step, classification based on allele frequency thresholds used data from the 1000 Genomes Project, gnomAD v4.1 exome and gnomAD v4.1 genome: BA1 (>5%), BS1 (1–5%), and PM2 (<0.05%). Next, computational predictions from SIFT, PolyPhen-2, and CADD assigned PP3 if at least two predicted deleterious effects, or BP4 if two predicted tolerated effects. The third step involved evidence from ClinVar for PP5 (pathogenic) and BP6 (benign) classifications. PM1 was applied when variants occurred within critical protein domains identified from Pfam. In the fourth step, literature evidence was integrated: PS3/BS3 (functional assays), PS4/BP5 (case-control studies), PP1/BS4 (cosegregation analysis), PS2/PM6 (confirmed or assumed de novo variants), and PP4 (clinical phenotyping supporting variant-disease association). Additional ACMG criteria were considered: BP7 (silent changes), PS1 and PM5 (same or different amino acid change at the same residue), PM3 and BP2 (compound heterozygosity in trans or cis), PM4 (indels outside repeat regions), and BP3 (indels within repeats). All evidence was compiled and interpreted using the University of Maryland’s Genetic Variant Interpretation Tool [23].

The PVS1 criterion was assigned to a loss-of-function (LoF) variant only if it satisfied five specific conditions. First, the variant had to be located in the major transcript of the gene, as defined by MANE. Second, the gene needed to be reported in ClinVar with at least 20 pathogenic or likely pathogenic (P/LP) LoF variants, contributing at least 5% of all the P/LP variants reported in that gene (that is, the variant count should be at least 20, contributing ≥5% of the total P/LP variants). Third, the variant had to be predicted as deleterious by AutoPVS1, either classified as ‘definitive’ in ClinGen or as ‘high confidence’ in SFARI. Fourth, the gene should have published literature supporting its association with autism spectrum disorder (ASD) or related overlapping clinical conditions involving a loss-of-function mechanism. Finally, any variant showing a relatively high minor allele frequency (MAF ≥ 1%) in the IndiGen dataset was excluded from PVS1 assignment.

### IndiGenomes vs Global: Frequency Comparison of Pathogenic Variants

For comparative analysis, ASD gene-panel-based variants classified as P/LP according to ACMG-AMP standards were considered. Minor allele frequencies (MAFs) of these variants were then extracted from the gnomAD database version 4.1.0 for major global populations represented by the following abbreviations: AFR (African/African American), AMR (Latino/Admixed American), ASJ (Ashkenazi Jewish), EAS (East Asian), FIN (Finnish), NFE (Non-Finnish European), SAS (South Asian), AMI (Amish), MID (Middle Eastern), and OTH (Other/Unassigned). Variant frequency comparisons across these populations were conducted, and statistical significance was determined using Fisher’s exact test with Bonferroni correction for multiple testing (p-value < 0.05).

## RESULTS

### Autism Spectrum Gene Prioritised from SFARI and ClinGen Databases

A total of 1,230 genes were retrieved from the SFARI database, and 329 genes from the ClinGen panel. The SFARI gene list included 304 Syndromic (S) genes, 234 High Confidence Score 1 (HC=1) genes, 711 strong candidate (SC=2) genes, and 191 suggestive evidence (SE=3) genes. The ClinGen panel comprised 254 genes in the Definitive (DE) category, 2 genes in the Strong category, 17 genes in the Moderate category, and 47 genes in the Other category. To generate a stringent ASD gene panel, genes from SFARI with an EAGLE score greater than 12 were combined with ClinGen genes classified as Definitive and Strong, resulting in a filtered set of genes. Additionally, SFARI genes categorised as Syndromic (S) and HC1 were integrated with ClinGen’s DE and Strong genes. After merging and removing duplicates, a final unique list of 517 ASD-associated genes was prepared for downstream analysis. The details of the stringent gene panel and its constituent genes are summarised in Figure 1 and Supplementary Table 1.

### Genetic Spectrum of ASD Related Gene Variants

A total of 1,798,502 variants were identified across ∼88 Mb of cumulative genomic regions spanning 517 ASD-associated genes, based on the gene panel selected for analysis. Following the initial filtering, 1,531,327 variants were retained with a minor allele frequency (MAF) < 5%, eliminating common variants and enriching the dataset for rare variants potentially relevant to ASD.

To assess missense variant pathogenicity, REVEL scores were used as a predictive filter. Applying a threshold of REVEL greater than 0.75, indicative of a high likelihood of deleterious impact, yielded 298 missense variants across 130 genes. Parallelly, loss-of-function (LoF) variants were identified using Variant Effect Predictor (VEP) in conjunction with the LoFTEE plugin. Initial annotation identified 584 LoF variants. High-confidence filtering was then applied by: (i) restricting to MANE Select transcripts, and (ii) retaining only those LoF variants flagged as high-confidence (HC) by LoFTEE. This filtering resulted in a refined set of 537 high-confidence LoF variants.

A total of 835 prioritised variants, including 298 missense and 537 loss-of-function (LoF) variants, were identified across 264 ASD-associated genes. Among these, 67 genes carried both missense and LoF variants, underscoring the broad spectrum of potentially impactful genetic alterations observed within ASD-associated genes. These variants represent a comprehensive set of potentially pathogenic alterations in ASD-associated genes within the Indian control population. The distribution and composition of these prioritised variants are detailed in Supplementary Table 2. The final list of predicted damaging variants based on the ASD gene panel is summarised in Figure 2, offering a visual overview of the genetic landscape.

**Figure 2:**
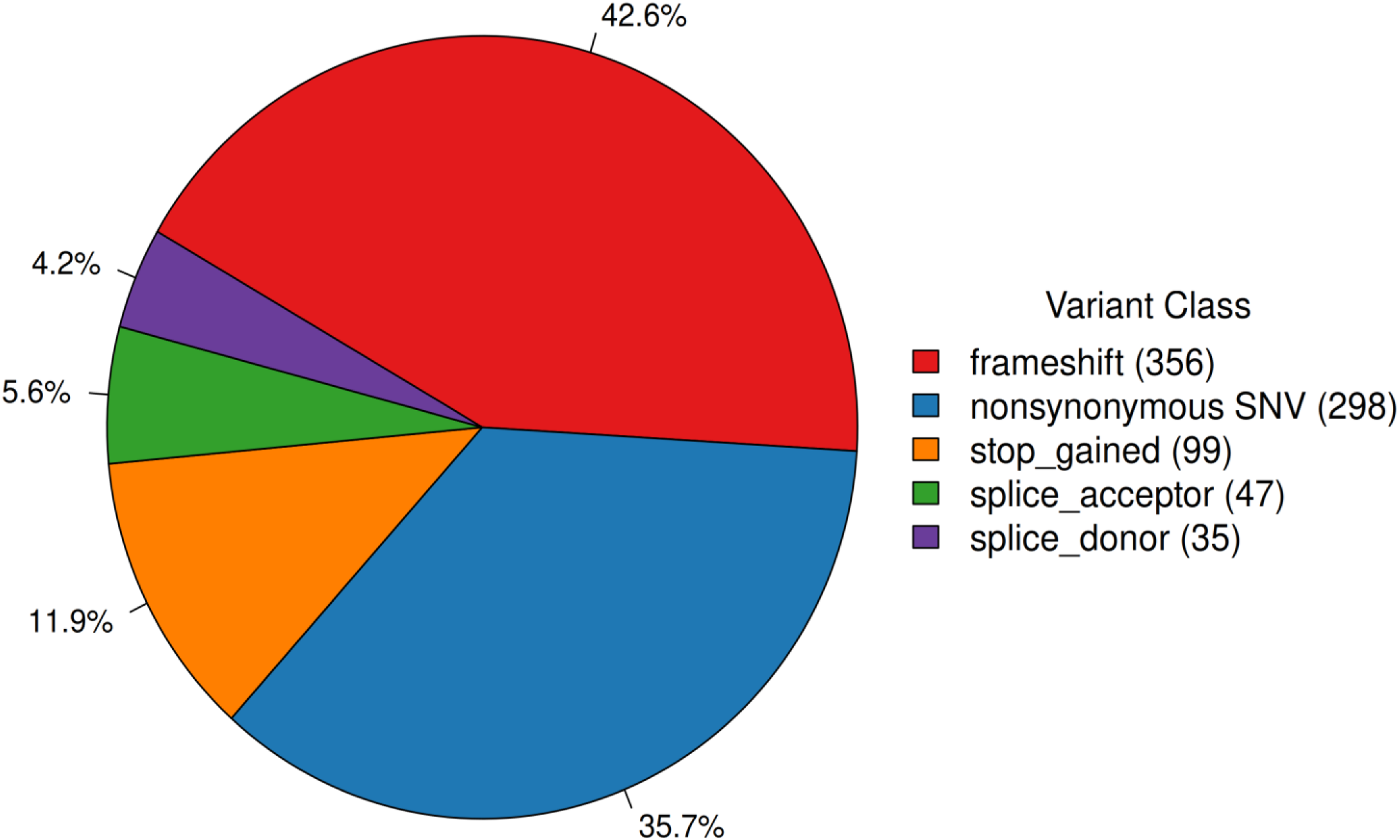
Percentage distribution of genetic variants across 517 genes in the IndiGenomes control dataset after variant prioritisation. The figure illustrates the proportion of different variant types or classes observed within the prioritised gene set.

### Assessment of Non-Synonymous/Missense Variants

In this analysis, 298 missense variants across 130 ASD-associated genes were assessed using ACMG-AMP classification criteria to evaluate their potential clinical significance. Among these, 1 variant was classified as pathogenic and 11 as likely pathogenic, collectively spanning 10 genes.

Additionally, 3 variants were likely benign, each found in a distinct gene. Notably, the majority, 283 variants across 128 genes, were classified as variants of uncertain significance (VUS), reflecting the current limitations in variant interpretation. The classification distribution, including pathogenic, likely pathogenic, likely benign, and VUS categories, is illustrated in Figure 3 as a stacked bar plot for visual clarity.

**Figure 3:**
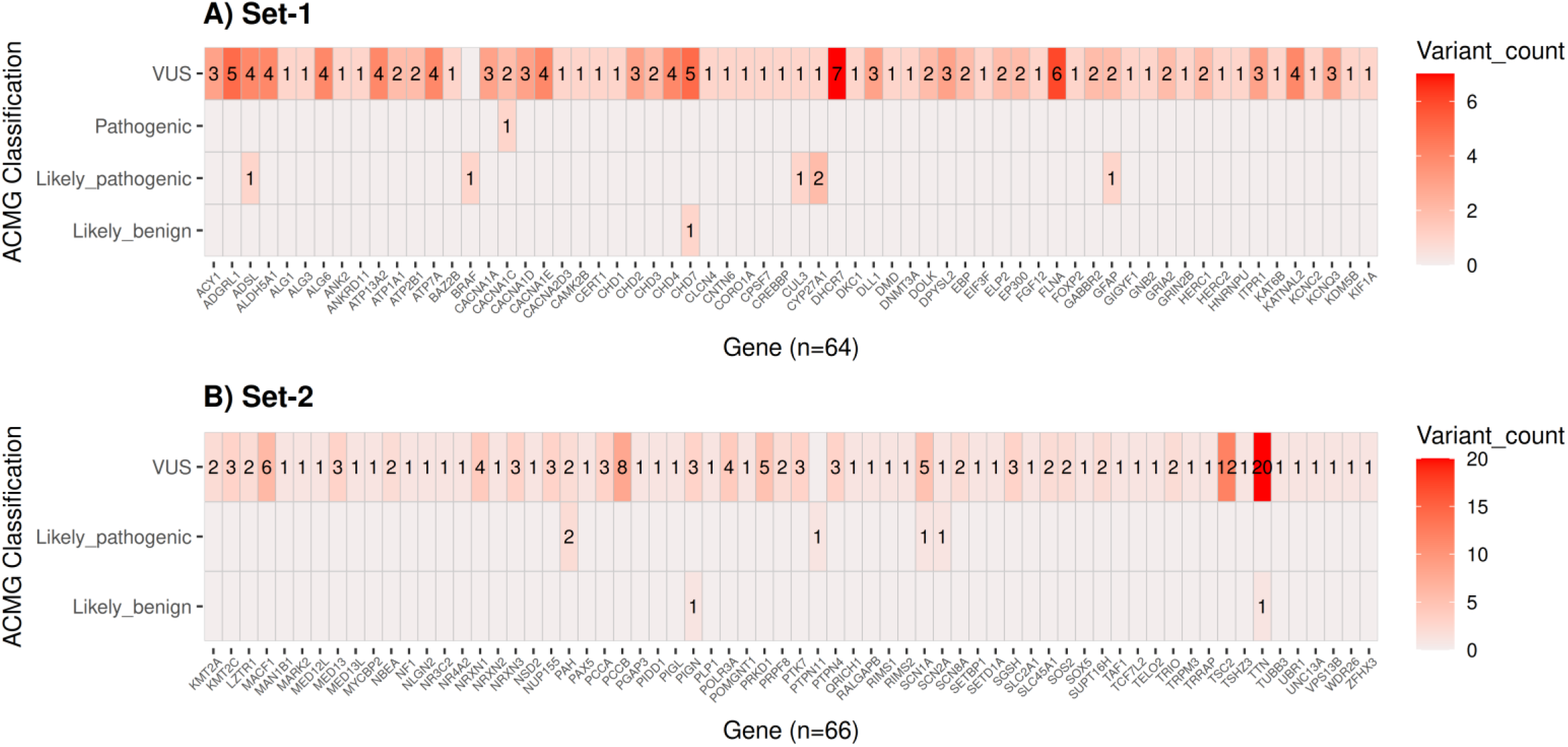
Distribution of classified non-synonymous variants based on ACMG guidelines in the ASD panel gene list. The figure includes two panels, Set-1 and Set-2, each showing the number of variants per gene categorised by ACMG classification (Pathogenic, Likely Pathogenic, VUS, and Likely Benign). This representation highlights the gene-wise burden of clinically relevant variants across the IndiGenomes dataset.

### Evaluation of Loss-of-Function (LoF) Variants

A total of 537 loss-of-function (LoF) variants were analysed and classified according to ACMG-AMP guidelines. This dataset included 47 splice-acceptor variants, 35 splice-donor variants, 356 frameshift insertions/deletions (indels), and 99 stop-gain variants. These were distributed across 201 ASD-associated genes, of which 18 genes harboured 43 variants classified as pathogenic. The remaining 494 variants across 187 genes were designated as variants of uncertain significance (VUS). The distribution of pathogenic variants across these 18 genes is depicted in Figure 4, showcasing the spectrum of high-confidence LoF variants and their ACMG classifications.

**Figure 4:**
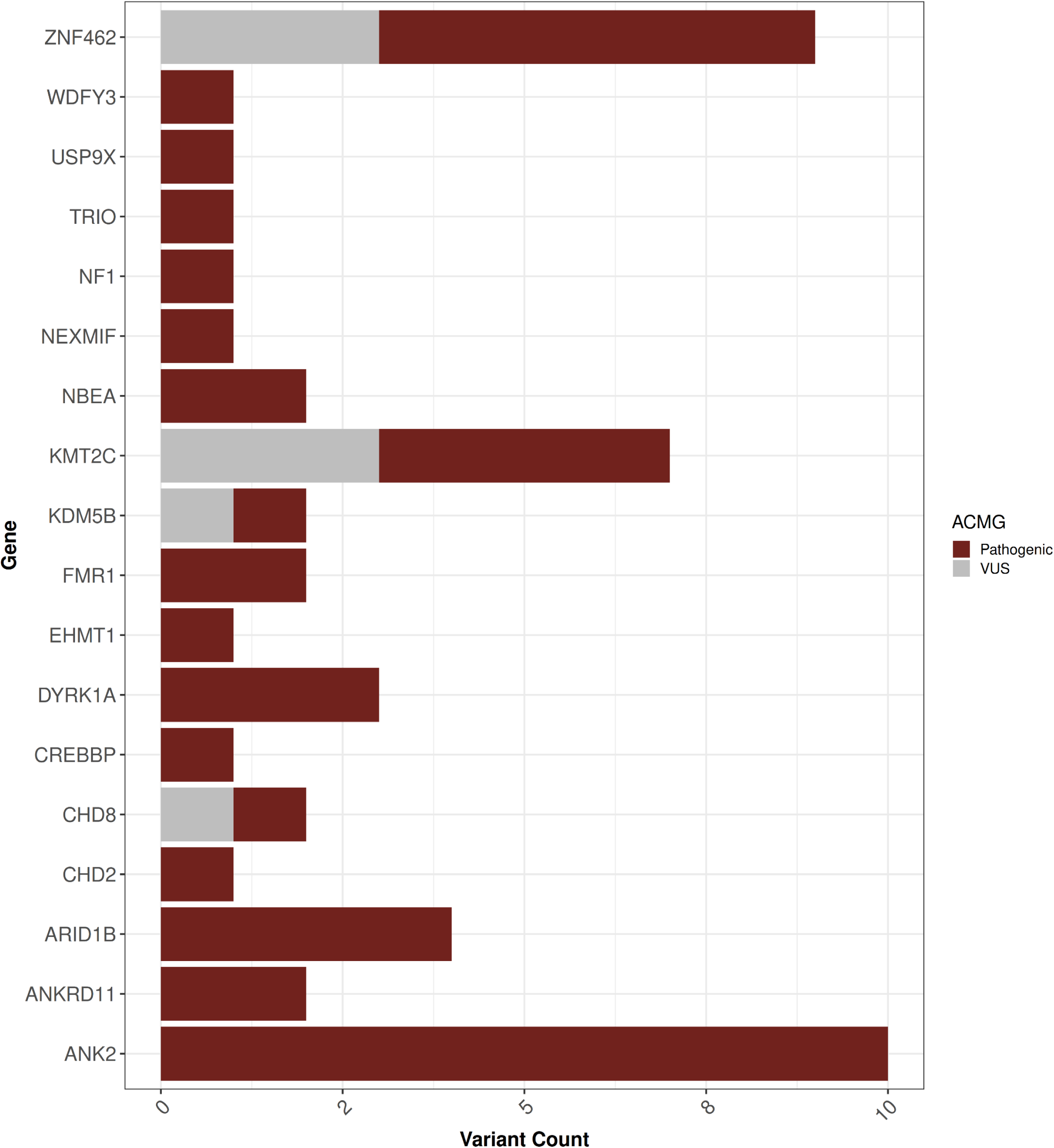
Depiction of all variants in genes containing Pathogenic (P) and Likely Pathogenic (LP) variants prioritised based on loss-of-function (LoF) impact from the ASD panel gene list. The stacked bar plot illustrates the number of LP and P variants across 19 genes harbouring LoF variants.

### ACMG Attributes Matrix of Classified Pathogenic and Likely Pathogenic Variants

According to the ACMG-AMP guidelines, all sorted, filtered, and prioritised variants, including missense and loss-of-function (LoF) variants, were systematically organised. A total of 55 variants were classified, comprising 44 pathogenic (P) and 11 likely pathogenic (LP) variants. These are visualised in Figure 5, where each variant is shown along the x-axis and the corresponding ACMG evidence attributes are plotted on the y-axis in a binary matrix. The presence of an ACMG attribute is marked as ’1’, and its absence is left blank. The matrix allows intuitive visualisation of the supporting evidence for each variant, with colour-coded bars indicating final classifications (pathogenic or likely pathogenic) based on ACMG criteria.

**Figure 5:**
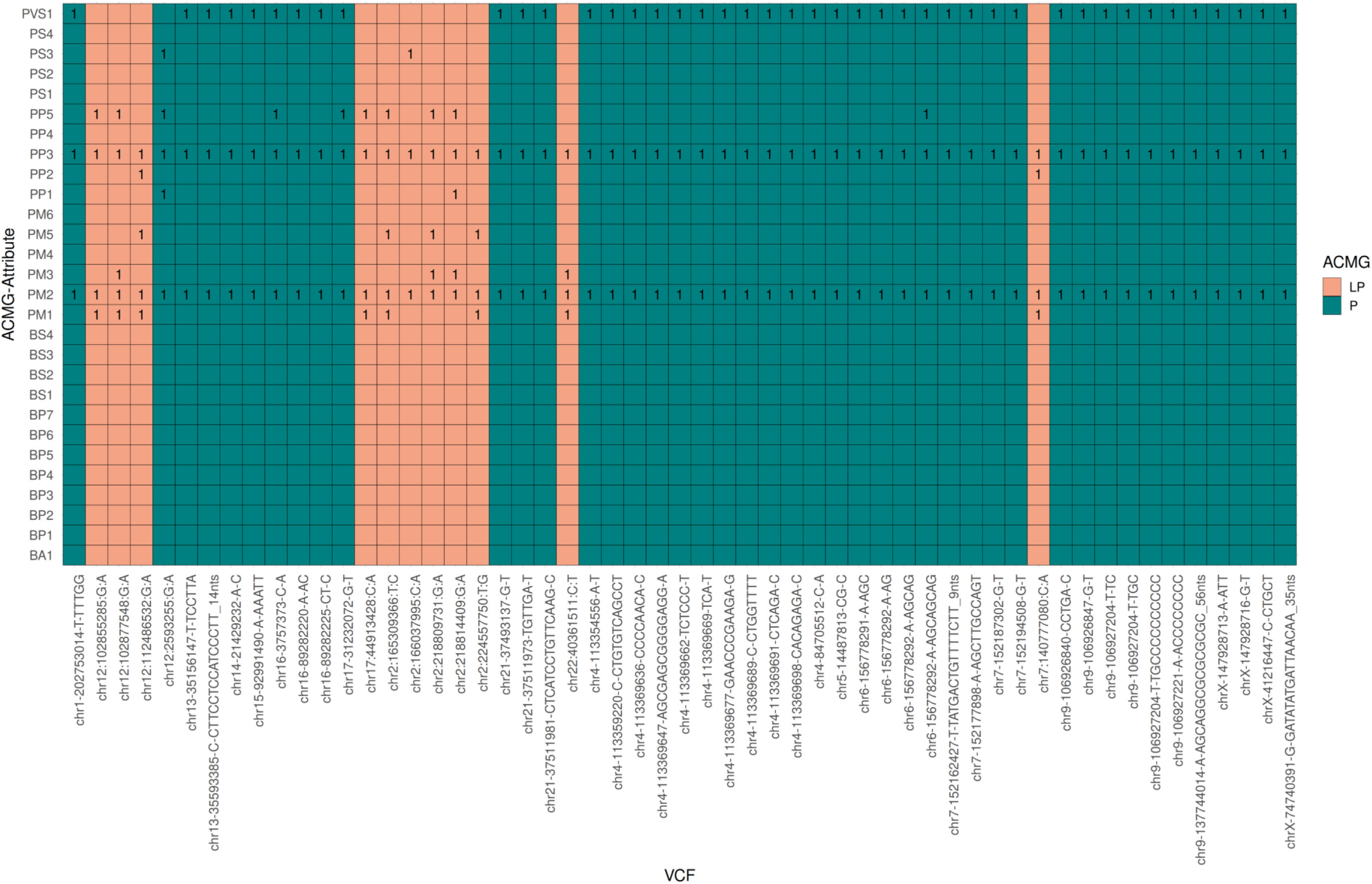
ACMG classification and supporting evidence for all Likely Pathogenic (LP) and Pathogenic (P) variants. The heatmap displays the presence of ACMG attributes used for classification across LP and P variants identified in the ASD panel gene list from the IndiGenomes control dataset.

### Zygosity Profiles and Inheritance Correlation of Variants in IndiGenome

To understand the individual-level genotype of 55 variants, LP (likely pathogenic) and P (Pathogenic) in the IndiGenome dataset, we investigated the homozygous (hom), heterozygous (het), and total allele count (AC) called across the IndiGenome dataset for a total of 28 genes. The thorough literature investigation of all the genes revealed that 3 genes follow the autosomal recessive (AR), 25 genes are autosomal dominant with conflicting evidence of mode of inheritance (AD), and 3 genes are X-linked with conflicting mode of inheritance (MOI). This analysis yielded an interesting observation regarding the zygosity of gene variants with MOI in the context of carriers and the anticipated genetic backgrounds of individuals with ASD. We identified 11 (1 in 93; 1.06%) expected carriers with a predicted genetic predisposition to ASD in control data, considering certain limitations of MOI, such as conflicting MOI and variability in disease penetrance. The complete set of analysed information is presented in Figure 6 and Table 1.

**Figure 6:**
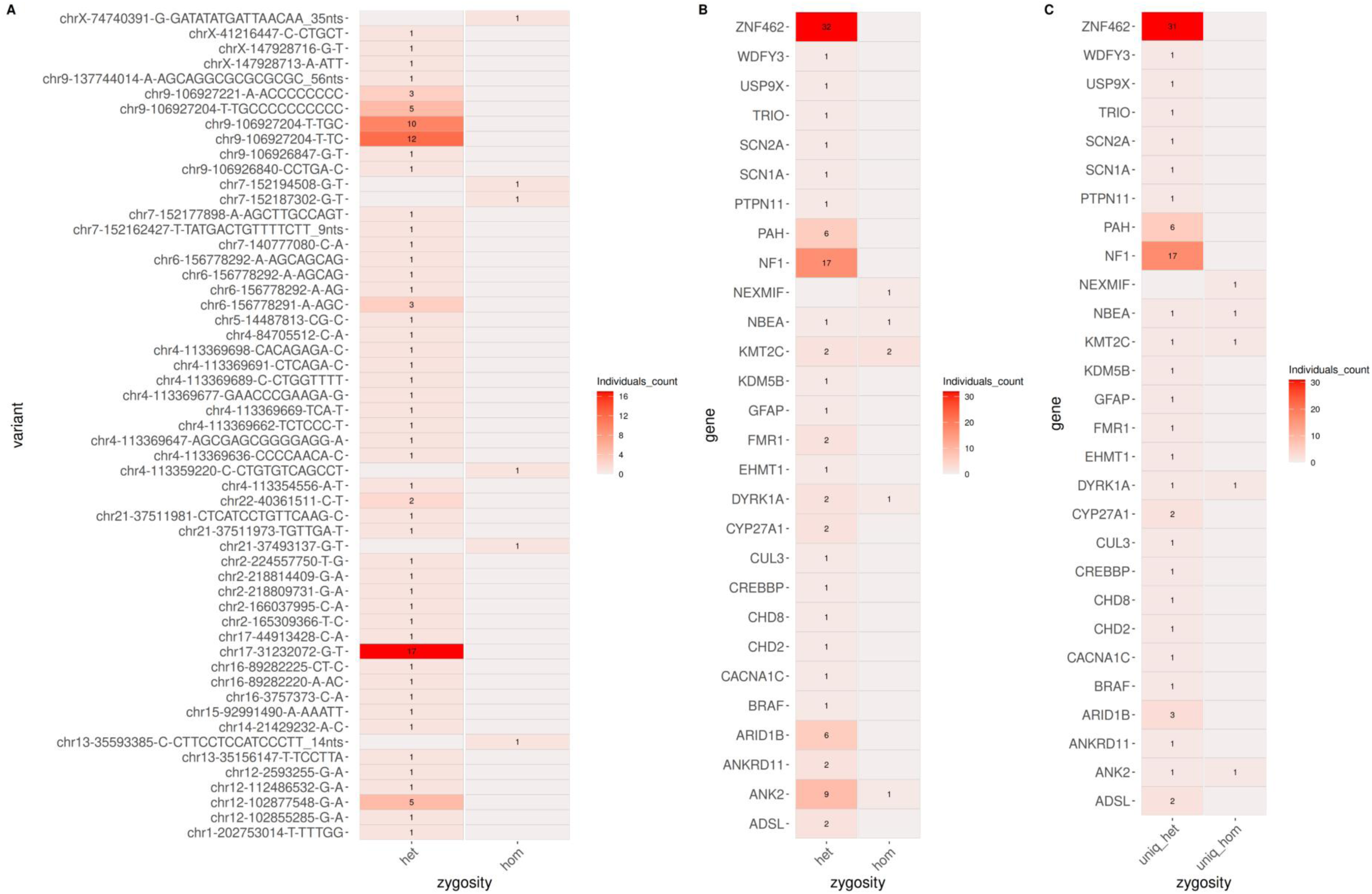
Zygosity distribution of ACMG-classified variants in the ASD gene panel. **(A)** Heatmap illustrating the counts of individuals with homozygous (hom) and heterozygous (het) Likely Pathogenic (LP) and Pathogenic (P) variants. **(B)** Heatmap showing the zygosity counts (hom and het) of LP and P variants stratified by gene. **(C)** Heatmap depicting counts of unique individuals with hom or het LP/P variants, where multiple occurrences of the same zygosity state within an individual are counted only once.

**Table 1:**
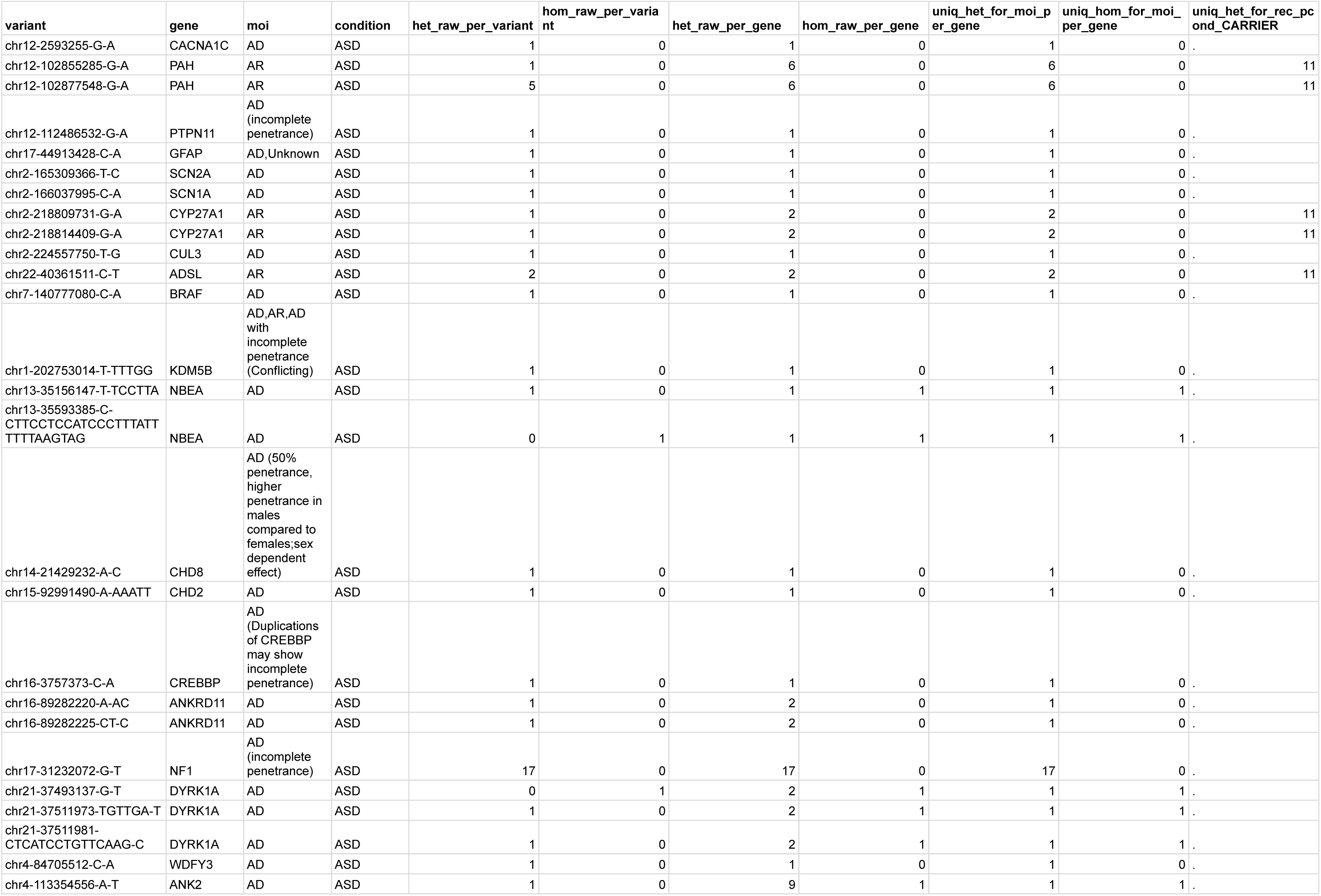
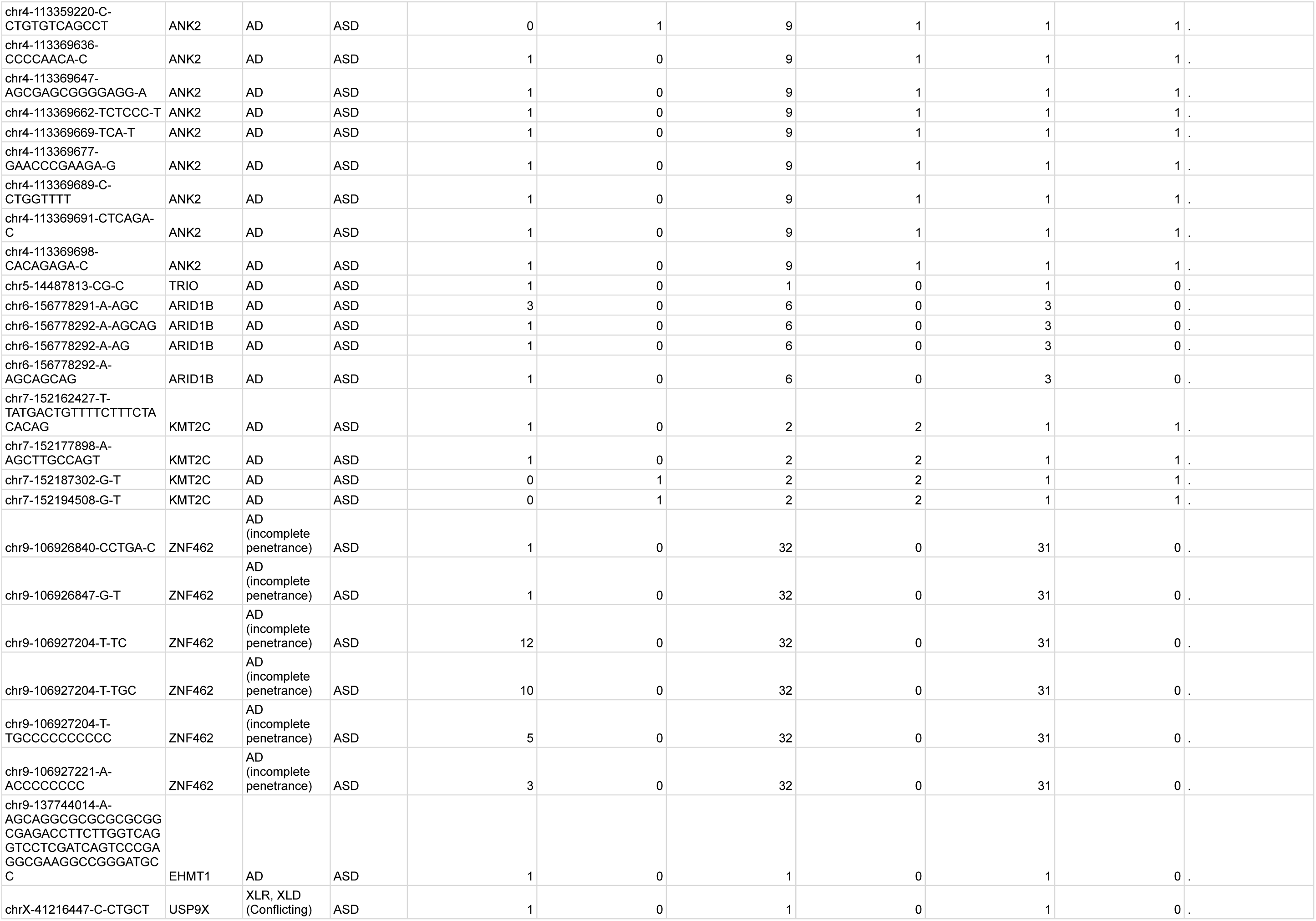
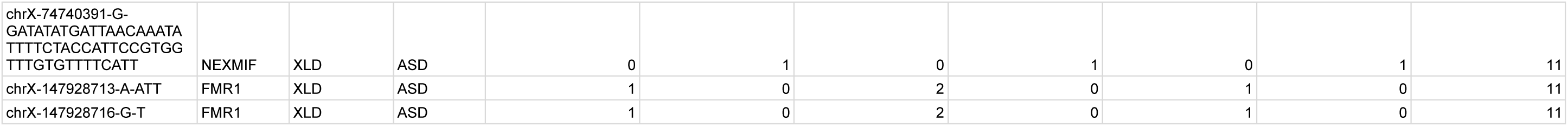
Haplotype profile of likely pathogenic (LP) and pathogenic (P) variants in the IndiGen control dataset. Notations: het_raw_per_variant = number of heterozygous genotypes per variant, hom_raw_per_variant = number of homozygous genotypes per variant, het_raw_per_gene = number of heterozygous genotypes per gene, hom_raw_per_gene = number of homozygous genotypes per gene, uniq_het_for_moi_per_gene = unique heterozygous variants per gene based on mode of inheritance (MOI) for the condition (ASD), uniq_hom_for_moi_per_gene = unique homozygous variants per gene based on MOI for the condition (ASD) and uniq_het_for_rec_pcond_CARRIER = unique heterozygous variants for recessive MOI per condition (ASD) representing carriers.

### Global Allele Frequency Analysis of ASD Gene Variants

A total of 55 genetic variants, 11 likely pathogenic and 44 pathogenic based on ACMG guidelines across 28 genes, were analysed to explore population-specific associations with Autism Spectrum Disorder (ASD). Minor Allele Frequencies (MAF) from the IndiGen dataset were compared with those from global populations. This comparison revealed 10 variants across 6 genes that are significantly enriched in the Indian population, using a corrected p-value threshold of less than 0.05. These enriched variants are visualised in a bubble plot, where they are encircled, with the corresponding genes prominently highlighted at the top of the graph (Figure 7). This population-specific enrichment underscores the importance of including diverse genetic backgrounds in ASD research for improved diagnosis and understanding of genetic risk in underrepresented groups.

**Figure 7:**
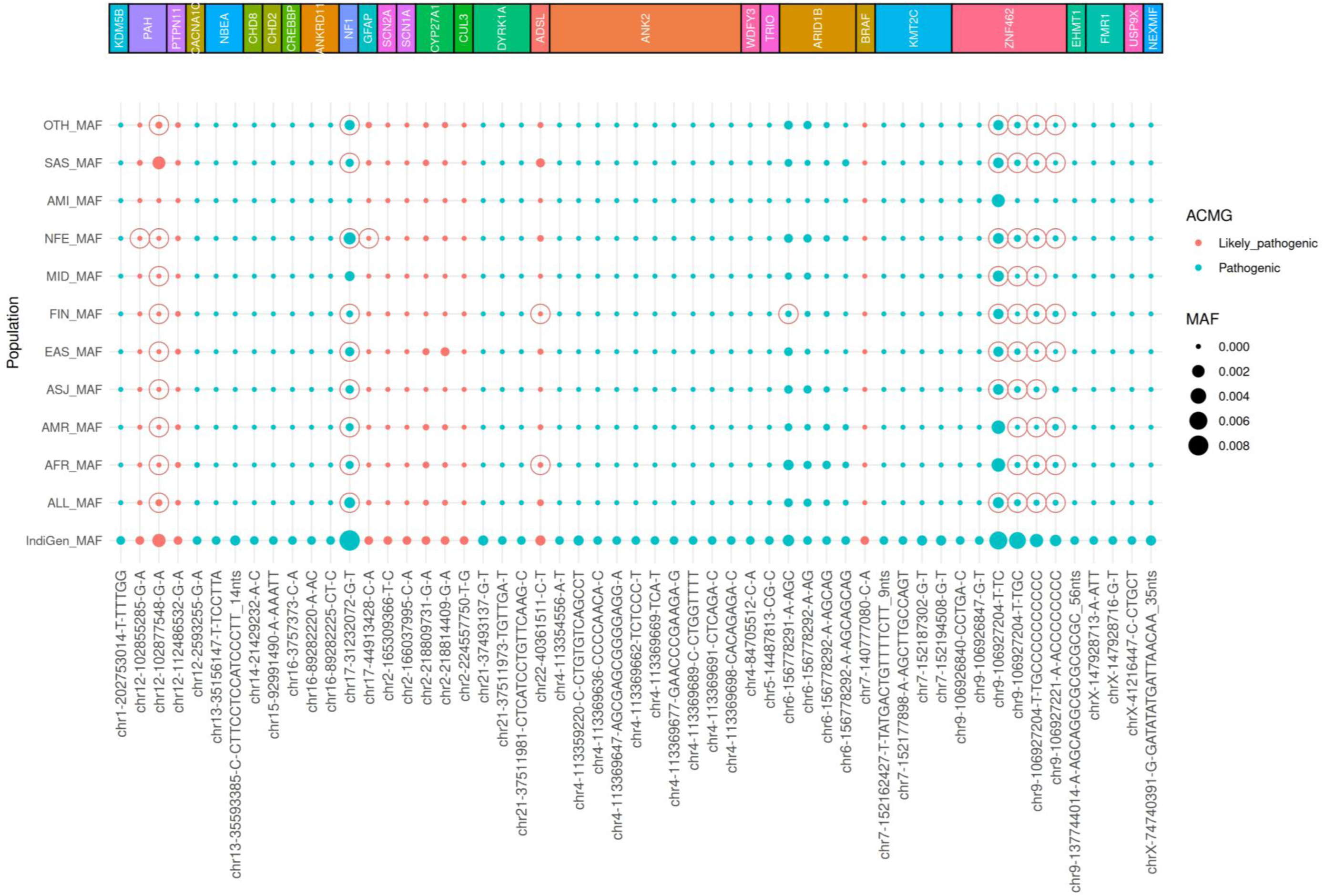
Cross-population comparison of minor allele frequencies (MAF) for Likely Pathogenic (LP) and Pathogenic (P) variants from IndiGenomes. This bubble plot visualises the MAF of LP and P variants identified in the IndiGenomes control dataset across major gnomAD v4.1 populations, including: African/African-American (AFR), Latino/Admixed American (AMR), Amish (AMI), Ashkenazi Jewish (ASJ), East Asian (EAS), Finnish (FIN), Non-Finnish European (NFE), Middle Eastern (MID), South Asian (SAS), and the Overall population. Variants are plotted on the x-axis, grouped by their respective genes, indicated at the top of the plot with an overlay graphic. Bubble size reflects the MAF in each population. Red-circled data points indicate statistically significant allele frequency differences (p < 0.05) based on Fisher’s exact test with Bonferroni correction.

## Discussion

This study offers the first insight into the genetic landscape and potential predisposition to Autism Spectrum Disorder (ASD) in an Indian control cohort, contributing to our broader understanding of its molecular basis in underrepresented populations. We analysed 1,798,502 variants from 517 genes with strong evidence of ASD association, sourced from the IndiGenomes dataset. Through stringent filtering, we prioritised 844 potentially damaging variants. Missense variants were filtered using a high-level confidence REVEL score greater than 0.75, while LoF variants were filtered using LoFTEE’s high-confidence (HC) designation[19,20]. Pathogenicity assessment using ACMG guidelines identified 55 variants composed of 44 pathogenic (P) and 11 likely pathogenic (LP) that include 12 missense from 10 genes and 43 loss-of-function (LoF) variants from 18 genes.

This concealed the finding with a higher burden observed in the LoF category in our analysis. Our findings underscore the importance of population-specific variant interpretation and lay the groundwork for future ASD-focused genetic studies in Indian populations.

The data from 55 P/LP variants, as interpreted by ACMG in 28 ASD-associated genes, suggest that 10 variants have been observed in other populations, as depicted in Figure 7. Literature evidence indicates that PAH, NBEA, ANKRD11, CYP27A1, and FMR1 each have atleast 2 P/LP variants linked to ASD, including biallelic, loss-of-function, truncating, and repeat-expansion mutations [24–28]. Similarly, DYRK1A, ANK2, ARID1B, KMT2C, and ZNF462 harbour atleast 3 or more such variants, spanning loss-of-function, rare damaging, and protein-truncating alterations associated with ASD [29–33]. Importantly, the analysis revealed LP/P variants in 13 genes from the 28-gene ASD panel (KDM5B, CHD8, CHD2, CREBBP, ANKRD11, DYRK1A, ANK2, ARID1B, KMT2C, CACNA1C, GFAP, SCN2A, and SCN1A), each previously present in a large ASD cohort study [34]. The recurrence of these genes and variant classes across studies not only reinforces their clinical relevance but also emphasises the value of integrating population-specific genomic data with global literature to refine diagnostic panels and improve risk assessment in ASD.

Genotyping of control data at the individual level was used to assess the distributions of homozygous and heterozygous variants across the 28 genes. Literature-supported classification of these genes by mode of inheritance (MOI), autosomal recessive (AR), autosomal dominant (AD), X-linked recessive (XLR), and X-linked dominant (XLD), revealed notable patterns in variant zygosity consistent with established MOIs, offering insights into carrier status and genetic backgrounds in the control population (Table 1 and Table 2). We identified 11 carrier individuals with genotypes potentially linked to ASD, while noting limitations such as uncertain MOI annotations and incomplete penetrance of certain genes (Figure 6, Table 1). Many ASD-associated genes containing pathogenic or likely pathogenic variants are reported to follow an autosomal dominant MOI, often exhibiting variable penetrance, incomplete dominance, and differing degrees of heritability. In this study, the control dataset comprised individuals already designated as healthy controls; therefore, the analysis was not based on their clinical disease status, as penetrance and heritability can vary between individuals [10,11]. This approach offers a comprehensive perspective on LP/P variants classified and organised according to rigorously standardised ACMG guidelines.

**Table 2:** Genes Implicated in Autism Spectrum Disorder: Modes of Inheritance (MOI), Phenotypic Spectrum, and Evidence from Literature.

|  |  |  |  |  |
| --- | --- | --- | --- | --- |
| **Gene** | **Inheritance** | **Known Phenotype for MOI** | **Literature Evidence Linking Genes to ASD** | **Evidence Based Comments** |
| KDM5B | AD,AR,AD with incomplete penetrance (Conflicting) | Autism | PMID: 39202393,PMID: 35905858 | KDM5B is mainly linked to autosomal recessive ID65, but heterozygous variants, both inherited and de novo have been reported in ASD and ID with variable expressivity. Inherited LoF variants often show incomplete penetrance, while de novo variants may have stronger pathogenic potential. |
| NBEA | AD | Neurodevelopment disorder with seizures | PMID: 30269351 | NBEA is a strong autism candidate gene, with de novo variants, monoallelic deletions, and chromosomal disruptions reported in individuals with ASD. It has also been linked to ID/DD |
| CHD8 | AD (50% penetrance, higher penetrance in males compared to females;sex dependent effect) | Autism | PMID: 32309624,PMID: 34440307,OMIM:* 610528 | CHD8 de novo variants are strongly associated with ASD, showing ~50% penetrance higher in males than females indicating a sex-dependent effect in autism risk. |
| CHD2 | AD | Epileptic encephalopathy | PMID: 35386198 | Mutations in the CHD2 gene are inherited in an autosomal-dominant manner and can lead to intellectual disability, epilepsy, and autism. |
| CREBBP | AD (Duplications of CREBBP may show incomplete penetrance) | Rubinstein-Taybi syndrome | PMID: 26730956,PMID: 19833603 | Mutations and duplications in CREBBP and EP300, key epigenetic regulators, are linked to ASD and developmental delays with variable features. |
| ANKRD11 | AD | KBG syndrome | PMID: 33262785 | A novel heterozygous ANKRD11 mutation in KBG syndrome was associated with severe intellectual disability, epilepsy, and autism-like features. |
| NF1 | AD (incomplete penetrance) | Neurofibromatosis-Noonan syndrome | PMID: 29204259,PMID: 34983638 | NF1, inherited in an autosomal dominant manner, has both inherited and de novo variants linked to ASD, with a prevalence of 11–26% in affected children compared to 1–4% in the general population, making it an important model for understanding autism's neurobiological basis. |
| DYRK1A | AD | Intellectual developmental disorder, autosomal dominant type 7 | PMID: 25707398,PMID: 37497568 | Disruptive de novo mutations in DYRK1A cause DYRK1A syndrome, a syndromic form of autism and intellectual disability, with 85% of affected individuals having ASD and 89% having ID, characterized by social communication deficits and sensory-seeking behaviors similar to idiopathic ASD with low IQ. |
| WDFY3 | AD | Primary Microcephaly or macrocephaly with developmental delay | PMID: 39614649,PMID: 34130600 | WDFY3 haploinsufficiency is linked to autosomal dominant neurodevelopmental disorders, macrocephaly, and autism susceptibility through its role in regulating Wnt signaling essential for neurodevelopment. |
| ANK2 | AD | neurodevelopmental disorder | PMID: 40035441 | ANK2 gene mutations are associated with autism spectrum disorder, epilepsy, and related neurodevelopmental issues, including language delay, intellectual disability, behavioral abnormalities, and ADHD, with a novel variant (p.R1003*) expanding the known genetic spectrum. |
| TRIO | AD | Intellectual disability | PMID: 26721934 | De novo TRIO loss-of-function mutations were found in individuals with mild to borderline ID and autistic or behavioral problems, with functional studies showing TRIO deficiency disrupts neurite outgrowth and synapse development. |

|  |  |  |  |  |  |
| --- | --- | --- | --- | --- | --- |
| ARID1B | AD | Coffin-Siris syndrome | PMID 22495309, PMID 23160955, PMID 21448237, PMID 21801163 | De novo disruptive variants, including frameshifts, deletions, and translocations in ARID1B, have been reported in multiple ASD patients, implicating it as an autism risk gene. | . |
| KMT2C | AD | Kleefstra syndrome 2 | Peduto A et al, 2023 | A child with developmental delay and autism was found to have a novel de novo KMT2C mutation causing Kleefstra syndrome type 2, showing typical features plus orthopedic and neurological anomalies. | . |
| ZNF462 | AD (incomplete penetrance) | Autistic behavior, Intellectual disability, Global developmental delay, | PMID: 31361404, PMID: 40538467, PMID: 31361404 | Weiss-Kruszka syndrome, caused by pathogenic ZNF462 variants, is an autosomal dominant neurodevelopmental disorder with variable expressivity and occasional incomplete penetrance, and ZNF462 loss-of-function variants can lead to a multiple congenital anomaly syndrome in which about one-third of affected individuals have autism spectrum disorder. | . |
| EHMT1 | AD | Kleefstra syndrome 2 | PMID 15805155, PMID 16826528 and PMID 19264732 | Disruptions in the EHMT1 gene are the cause of Kleefstra syndrome, which is accompanied by autistic features and intellectual disability. | . |
| USP9X | XLR, XLD (Conflicting) | Intellectual developmental disorder-X-linked 99, Intellectual developmental disorder-X-linked 99-syndromic-female-restricted | PMID: 31443933 | USP9X variants cause a distinctive neurodevelopmental and behavioral syndrome in males, characterized by brain structural abnormalities, global developmental delay with speech, language, and behavioral alterations, and are linked to disrupted TGF-β signaling and hippocampal function. | . |
| NEXMIF | XLD | Intellectual disability | PMID: 23615299 | KIAA2022 deficiency causes X-linked intellectual disability with autistic features by impairing neurite outgrowth and disrupting brain development. | . |
| FMR1 | XLD | Fragile X syndrome | PMID: 11773805, PMID: 16700053, PMID: 16700053 | This study found that one-third of young children with Fragile X syndrome show autism-like behaviors, suggesting possible extra genetic factors. | . |
| CACNA1C | AD | Timothy syndrome, | PMID: 15454078 | Timothy syndrome, caused by a de novo Ca(V)1.2 (CACNA1C, α1C, α11.2) G406R mutation, leads to multiorgan dysfunction including lethal arrhythmias, congenital anomalies, and autism, through impaired channel inactivation and Ca<sup>2+</sup> overload. | . |
| PAH | AR | Non-phenylketonuria hyperphenylalaninemia | PMID: 34121999 | Two consanguineous families with homozygous nonsense or frameshift mutations in PAH, the gene responsible for phenylketonuria, a well-known early neurometabolic cause of ASD, were clinically confirmed to have phenylketonuria. | . |
| PTPN11 | AD (incomplete penetrance) | Noonan syndrome | PMID: 11704759, PMID: 28191889 | Rare PTPN11 mutations, causing autosomal dominant Noonan syndrome, are linked to syndromic autism in a subset of affected individuals. | . |
| GFAP | AD, Unknown | Alexander disease, Autism | PMID: 23840007 | Elevated brain sAPPα levels in some individuals with autism are linked to increased GFAP expression, gliosis, and autism-like behaviors, as shown in both patient samples and transgenic mouse models. | . |
| SCN2A | AD | SCN2A-Related Disorders | PMID: 24650168, PMID: 23675382 | A de novo SCN2A splice site mutation in a boy with ASD likely causes protein loss, supporting its role in ASD with cognitive, social, and sensory impairments. | . |
| SCN1A | AD | SCN1A-Related seizure disorders | PMID: 30060894 | First report on the association of SCN1A mutation, childhood schizophrenia and autism spectrum disorder without epilepsy | . |
| CYP27A1 | AR | Cerebrotendinous xanthomatosis | PMID: 28894950 | ASD was found in 13% of CTX patients, often as an early feature before other symptoms. Screening for CTX is recommended in ASD cases with diarrhea, intellectual disability, cataract, or neurological signs, as early treatment improves outcomes. | . |
| CUL3 | AD | Neurodevelopmental disorder with or without autism or seizures | PMID 22495309,PMID 22914163 | A non-familial autistic proband had a de novo stopgain mutation (p.R546X) in CUL3, previously linked to hypertension and electrolyte issues; another autistic case carried a similar de novo stopgain (p.E246X). | . |
| ADSL | AR | Adenylosuccinase deficiency | Naghiloo et al, (2025) | A Turkish patient with epilepsy, autism spectrum symptoms, developmental delay, and seizures was found by WES to have two ADSL variants (p.R452C, p. R337Q), suggesting ADSL deficiency as the underlying cause. | . |
| BRAF | AD | Cardiofaciocutaneous syndrome and Noonan syndrome | PMID: 25363760,PMID: 33004838,PMID: 24458522,PMID: 24101678 | De novo missense variants in BRAF, enriched in ASD cohorts, are linked to cardiofaciocutaneous syndrome (AD inheritance), in which autistic features are frequently observed. | . |

Population-level analysis revealed 10 variants across six genes (PAH, NF1, ADSL, TRIO, and ZNF462) with significantly higher frequencies in the Indian population (corrected p < 0.05, Fisher’s exact test). Most of these variants showed concordance with minor allele frequencies reported in South Asian (SAS) populations. These enriched variants, illustrated in the bubble plot (Figure 7), highlight the importance of incorporating diverse and underrepresented genetic backgrounds into ASD research. Such inclusion can improve the diagnostic yield, refine variant interpretation, and enhance our understanding of population-specific genetic risk factors in ASD.

## Data Availability

All data produced in the present study are available upon reasonable request to the authors

## Acknowledgement

MK, TI, and MS acknowledge financial support from the project grant (Code: CLP009037). SS acknowledges the University Grants Commission, Senior Research Fellowship (UGC-SRF) for fellowship support. This research was additionally supported by funding from the Council of Scientific and Industrial Research (CSIR). Computational analyses were performed using the High-Performance Computing (HPC) facility at CSIR-IGIB.

## Authors Contribution

BK conceived the study, supervised the analysis, and proofread the manuscript. MK designed the data analysis pipeline, prepared the initial manuscript draft, interpreted the variants, and performed a thoroughly conceptualised analysis. TI carried out formal data analysis, interpreted variants, and proofread the manuscript. MS and SS conducted formal analysis, variant interpretation and proofread the manuscript. SG contributed valuable insights that informed and strengthened the study. SSB and VS provided genomic sequencing data and proofread the manuscript.

## Data Availability

The datasets supporting the findings of this study are available from the corresponding author upon reasonable request. However, the necessary supplementary data are provided with the manuscript as supplementary files.

## Statement of Ethics

The authors attest that they obtained all the necessary participant consent papers. The participant has provided his agreement in the form of his clinical information to be published in the journal. The participants understand that their names and initials will not be published.

## Supplementary data

**Supplementary Table 1:** Supplementary Table 1: Prioritisation of ASD gene panel, showing 517 unique genes from SFARI and ClinGen. Abbreviations: S-Syndromic, HC-High Confidence, SC-Strong Candidate, SE-Suggestive Evidence, DE-Definitive, 1S (HC+S),2S(SC+S), 3S(SE+S)

**Supplementary Table 2:** Predicted damaging genetic variants identified from the IndGenomes dataset using the ASD gene panel (517 genes), filtered using REVEL scores >0.75 for missense variants and LoFTEE-predicted high-confidence loss-of-function (LoF) variants.

**Supplementary Table 3:** Highlighting the PVS1 assignment criteria, the satisfying variants (P/LP) are listed with their parameters.

