## Supplementary Table 1-3 for "Genetic Spectrum of Autism Spectrum Disorder Associated Variants in the Indian Population: Insights from 1,029 IndiGenomes"

| **Supplementary Table 1:** Supplementary Table 1: Prioritisation of ASD gene panel, showing 517 unique genes from SFARI and ClinGen. Abbreviations: S-Syndromic, HC-High Confidence, SC-Strong Candidate, SE-Suggestive Evidence, DE-Definitive, 1S (HC+S),2S(SC+S), 3S(SE+S) | | | |
|---|---|---|---|
| **Gene** | **Sfari_Gene_Score; ClinGen_Class** | **set** | **Eagle Score; ClinGen_Class** |
| ACTB | 1S(HC+S) ; . | set-1 | . |
| ACTL6B | S; . | set-1 | . |
| ACY1 | S; . | set-1 | . |
| ADGRL1 | 3S(SE+S) ; . | set-1 | . |
| ADNP | 1S(HC+S) ; Definitive | set-1 | . |
| ADSL | 1S(HC+S) ; Definitive | set-1 | . |
| AGO2 | 2S(SC+S) ; . | set-1 | . |
| AHDC1 | 1S(HC+S) ; Definitive | set-1 | . |
| AHI1 | S; . | set-1 | . |
| ALDH1A3 | S; . | set-1 | . |
| ALDH5A1 | 1S(HC+S) ; Definitive | set-1 | . |
| ALG6 | S; Definitive | set-1 | . |
| ANKRD11 | 1S(HC+S) ; Definitive | set-1 | . |
| ANKRD17 | 2S(SC+S) ; . | set-1 | . |
| ANKS1B | 2S(SC+S) ; . | set-1 | . |
| AP1S2 | S; Definitive | set-1 | . |
| ARHGEF9 | 1S(HC+S) ; Definitive | set-1 | . |
| ARID1A | 3S(SE+S) ; Definitive | set-1 | . |
| ARID1B | 1S(HC+S) ; Definitive | set-1 | . |
| ARID2 | 2S(SC+S) ; Definitive | set-1 | . |
| ARX | 1S(HC+S) ; Definitive | set-1 | . |
| ASXL3 | 1S(HC+S) ; Definitive | set-1 | . |
| ATP1A1 | 2S(SC+S) ; . | set-1 | . |
| ATP1A3 | 2S(SC+S) ; . | set-1 | . |
| ATP2B1 | 3S(SE+S) ; . | set-1 | . |
| BCL11A | 1S(HC+S) ; Definitive | set-1 | . |
| BCORL1 | S; Limited | set-1 | . |
| BICRA | 2S(SC+S) ; . | set-1 | . |

| | | | |
|---|---|---|---|
| BRAF | 1S(HC+S) ; . | set-1 | . |
| BRSK2 | 1S(HC+S) ; Definitive | set-1 | . |
| BRWD3 | S; Definitive | set-1 | . |
| C12orf57 | S; . | set-1 | . |
| CACNA1A | 1S(HC+S) ; . | set-1 | . |
| CACNA1C | 1S(HC+S) ; . | set-1 | . |
| CAMK2A | 2S(SC+S) ; . | set-1 | . |
| CAMK2B | S; . | set-1 | . |
| CAMK2D | 3S(SE+S) ; . | set-1 | . |
| CBX1 | 3S(SE+S) ; . | set-1 | . |
| CCNK | S; . | set-1 | . |
| CDH2 | 3S(SE+S) ; . | set-1 | . |
| CDK13 | S; Definitive | set-1 | . |
| CDK19 | 3S(SE+S) ; . | set-1 | . |
| CDK8 | S; . | set-1 | . |
| CDKL5 | 1S(HC+S) ; . | set-1 | . |
| CELF2 | 2S(SC+S) ; . | set-1 | . |
| CEP290 | 2S(SC+S) ; . | set-1 | . |
| CERT1 | 3S(SE+S) ; . | set-1 | . |
| CHAMP1 | 1S(HC+S) ; Definitive | set-1 | . |
| CHD1 | 2S(SC+S) ; Limited | set-1 | . |
| CHD2 | 1S(HC+S) ; . | set-1 | . |
| CHD3 | 1S(HC+S) ; Definitive | set-1 | . |
| CHD7 | 1S(HC+S) ;. | set-1 | . |
| CHD8 | 1S(HC+S) ; Definitive | set-1 | . |
| CHD4 | 3S(SE+S) ; Definitive | set-1 | . |
| CHKB | S; . | set-1 | . |
| CLCN4 | 2S(SC+S) ; Definitive | set-1 | . |
| CNKSR2 | 2S(SC+S) ; Definitive | set-1 | . |
| CNOT1 | 2S(SC+S) ; Definitive | set-1 | . |
| CNOT3 | 1S(HC+S) ; Definitive | set-1 | . |

| | | | |
|---|---|---|---|
| CNTNAP2 | 2S(SC+S) ; Disputed | set-1 | . |
| CREBBP | 1S(HC+S) ; Definitive | set-1 | . |
| CSDE1 | 1S(HC+S) ; Definitive | set-1 | . |
| CSNK1G1 | 3S(SE+S) ; . | set-1 | . |
| CSNK2A1 | 1S(HC+S) ; Definitive | set-1 | . |
| CSNK2B | S; . | set-1 | . |
| CTCF | 1S(HC+S) ; Definitive | set-1 | . |
| CTNNA2 | S; . | set-1 | . |
| CTR9 | 3S(SE+S) ; . | set-1 | . |
| CUL4B | 3S(SE+S) ; Definitive | set-1 | . |
| CUX2 | 2S(SC+S) ; . | set-1 | . |
| CYP27A1 | S; . | set-1 | . |
| DDX23 | S; . | set-1 | . |
| DDX3X | 1S(HC+S) ; Definitive | set-1 | . |
| DEAF1 | 1S(HC+S) ; . | set-1 | . |
| DEPDC5 | S; . | set-1 | . |
| DHCR7 | 1S(HC+S) ; Definitive | set-1 | . |
| DHX30 | S; Definitive | set-1 | . |
| DHX9 | 3S(SE+S) ; . | set-1 | . |
| DLL1 | 2S(SC+S) ; . | set-1 | . |
| DMD | S; . | set-1 | . |
| DMPK | 1S(HC+S) ; . | set-1 | . |
| DNMT3A | 1S(HC+S) ; Limited | set-1 | . |
| DOLK | S; . | set-1 | . |
| DYRK1A | 1S(HC+S) ; Definitive | set-1 | . |
| EBF3 | 1S(HC+S) ; Definitive | set-1 | . |
| EEF1A2 | S; . | set-1 | . |
| EHMT1 | 1S(HC+S) ; Definitive | set-1 | . |
| ELP2 | 2S(SC+S) ; . | set-1 | . |
| EP300 | 1S(HC+S) ; . | set-1 | . |
| FBRSL1 | S; . | set-1 | . |

| | | | |
|---|---|---|---|
| FBXO11 | 2S(SC+S) ; . | set-1 | . |
| FGF13 | 3S(SE+S) ; . | set-1 | . |
| FMR1 | 1S(HC+S) ; Definitive | set-1 | . |
| FOXG1 | 1S(HC+S) ; . | set-1 | . |
| FOXP1 | 1S(HC+S) ; Definitive | set-1 | . |
| FRMD5 | 3S(SE+S) ; . | set-1 | . |
| FRMPD4 | S; Definitive | set-1 | . |
| FRYL | 3S(SE+S) ; . | set-1 | . |
| GABBR2 | 2S(SC+S) ; . | set-1 | . |
| GABRA3 | S; . | set-1 | . |
| GALNT2 | S; . | set-1 | . |
| GATM | S; . | set-1 | . |
| GNAI1 | 1S(HC+S) ; Definitive | set-1 | . |
| GNB2 | S; . | set-1 | . |
| GRIA3 | S; Definitive | set-1 | . |
| H1-4 | S; Definitive | set-1 | . |
| H3-3B | 3S(SE+S) ; . | set-1 | . |
| H4C11 | S; . | set-1 | . |
| H4C3 | S; . | set-1 | . |
| H4C5 | S; . | set-1 | . |
| HCFC1 | S; Definitive | set-1 | . |
| HDAC4 | 2S(SC+S) ; . | set-1 | . |
| HDAC8 | S; Definitive | set-1 | . |
| HEPACAM | S; Definitive | set-1 | . |
| HERC1 | 2S(SC+S) ; . | set-1 | . |
| HERC2 | S; . | set-1 | . |
| HIVEP2 | 1S(HC+S) ; . | set-1 | . |
| HNRNPD | 2S(SC+S) ; . | set-1 | . |
| HNRNPK | 2S(SC+S) ; Definitive | set-1 | . |
| HNRNPR | 2S(SC+S) ; Definitive | set-1 | . |
| HNRNPU | 1S(HC+S) ; Definitive | set-1 | . |

| | | | |
|---|---|---|---|
| HNRNPUL2 | 2S(SC+S) ; . | set-1 | . |
| HOXA1 | S; Definitive | set-1 | . |
| HUWE1 | S; Definitive | set-1 | . |
| INTS1 | S; . | set-1 | . |
| IQSEC2 | 1S(HC+S) ; Definitive | set-1 | . |
| IRF2BPL | 1S(HC+S) ; . | set-1 | . |
| IRX5 | 3S(SE+S) ; . | set-1 | . |
| KANSL1 | 1S(HC+S) ; Definitive | set-1 | . |
| KAT6A | 2S(SC+S) ; Definitive | set-1 | . |
| KCNB1 | 1S(HC+S) ; . | set-1 | . |
| KCNH1 | 3S(SE+S) ; Definitive | set-1 | . |
| KDM3B | 1S(HC+S) ; Definitive | set-1 | . |
| KIF1A | 2S(SC+S) ; Definitive | set-1 | . |
| KIF5C | 2S(SC+S) ; . | set-1 | . |
| KMT2A | 1S(HC+S) ; . | set-1 | . |
| KMT2C | 1S(HC+S) ; Definitive | set-1 | . |
| KMT2E | 1S(HC+S) ; Definitive | set-1 | . |
| KPTN | S; . | set-1 | . |
| LNPK | S; . | set-1 | . |
| MACF1 | 3S(SE+S) ; . | set-1 | . |
| MAGEL2 | 1S(HC+S) ; Definitive | set-1 | . |
| MBD5 | 1S(HC+S) ; Definitive | set-1 | . |
| MBOAT7 | 1S(HC+S) ; Definitive | set-1 | . |
| MECP2 | 1S(HC+S) ; . | set-1 | . |
| MED12L | 2S(SC+S) ; . | set-1 | . |
| MED13 | 1S(HC+S) ; Definitive | set-1 | . |
| MED13L | 1S(HC+S) ; Definitive | set-1 | . |
| MEF2C | 1S(HC+S) ; Definitive | set-1 | . |
| MEIS2 | 1S(HC+S) ; Definitive | set-1 | . |
| MSL3 | 1S(HC+S) ; Definitive | set-1 | . |
| MSX2 | 3S(SE+S) ; . | set-1 | . |

| | | | |
|---|---|---|---|
| MTOR | 1S(HC+S) ; . | set-1 | . |
| MTSS2 | S; . | set-1 | . |
| NAA10 | 3S(SE+S) ; Definitive | set-1 | . |
| NAA15 | 1S(HC+S) ; Definitive | set-1 | . |
| NACC1 | 1S(HC+S) ; Definitive | set-1 | . |
| NBEA | 1S(HC+S) ; Definitive | set-1 | . |
| NF1 | 1S(HC+S) ; . | set-1 | . |
| NFIB | 2S(SC+S) ; Definitive | set-1 | . |
| NFIX | S; . | set-1 | . |
| NIPBL | 1S(HC+S) ; Definitive | set-1 | . |
| NOVA2 | S; . | set-1 | . |
| NR2F1 | 2S(SC+S) ; Definitive | set-1 | . |
| NR3C2 | 1S(HC+S) ; . | set-1 | . |
| NSD1 | 1S(HC+S) ; Definitive | set-1 | . |
| NSD2 | 2S(SC+S) ; Definitive | set-1 | . |
| NTNG1 | 2S(SC+S) ; Limited | set-1 | . |
| NTNG2 | S; . | set-1 | . |
| NTRK2 | S; . | set-1 | . |
| OCRL | S; Definitive | set-1 | . |
| PABPC1 | 3S(SE+S) ; . | set-1 | . |
| PACS1 | 1S(HC+S) ; Definitive | set-1 | . |
| PACS2 | S; Definitive | set-1 | . |
| PAK1 | S; . | set-1 | . |
| PAX6 | S; . | set-1 | . |
| PCCA | S; . | set-1 | . |
| PCCB | 1S(HC+S) ; . | set-1 | . |
| PCDH19 | 1S(HC+S) ; . | set-1 | . |
| PDZD8 | S; . | set-1 | . |
| PHF21A | 1S(HC+S) ; Definitive | set-1 | . |
| PHF8 | S; Definitive | set-1 | . |
| PHIP | 1S(HC+S) ; Definitive | set-1 | . |

| | | | |
|---|---|---|---|
| PIK3R2 | S; . | set-1 | . |
| PJA1 | 3S(SE+S) ; . | set-1 | . |
| POGZ | 1S(HC+S) ; Definitive | set-1 | . |
| POLR2A | 3S(SE+S) ; . | set-1 | . |
| POLR3A | 3S(SE+S) ; . | set-1 | . |
| POMGNT1 | 1S(HC+S) ; . | set-1 | . |
| POU3F3 | S; Definitive | set-1 | . |
| PPFIA3 | 3S(SE+S) ; . | set-1 | . |
| PPM1D | 2S(SC+S) ; Definitive | set-1 | . |
| PPP2CA | S; . | set-1 | . |
| PPP2R5D | 1S(HC+S) ; Definitive | set-1 | . |
| PPP3CA | 3S(SE+S) ; . | set-1 | . |
| PRKD1 | S; . | set-1 | . |
| PRODH | 2S(SC+S) ; . | set-1 | . |
| PRPF8 | 3S(SE+S) ; . | set-1 | . |
| PRR12 | 1S(HC+S) ; . | set-1 | . |
| PSMD12 | 1S(HC+S) ; Definitive | set-1 | . |
| PPP2R5C | 3S(SE+S) ; . | set-1 | . |
| PTEN | 1S(HC+S) ; . | set-1 | . |
| PTPN11 | 1S(HC+S) ; . | set-1 | . |
| PTPN4 | S; . | set-1 | . |
| PUF60 | 3S(SE+S) ; Definitive | set-1 | . |
| RAC1 | S; Definitive | set-1 | . |
| RAD21 | S; Definitive | set-1 | . |
| RAI1 | 1S(HC+S) ; Definitive | set-1 | . |
| RALA | S; Definitive | set-1 | . |
| RERE | 1S(HC+S) ; Definitive | set-1 | . |
| RFX4 | 3S(SE+S) ; . | set-1 | . |
| RFX7 | 3S(SE+S) ; Definitive | set-1 | . |
| RHEB | S; . | set-1 | . |
| RIMS2 | 3S(SE+S) ; . | set-1 | . |

| | | | |
|---|---|---|---|
| RLIM | S; . | set-1 | . |
| RNF135 | 2S(SC+S) ; . | set-1 | . |
| RNU4-2 | 1S(HC+S) ; . | set-1 | . |
| RORA | S; . | set-1 | . |
| RORB | 1S(HC+S) ; . | set-1 | . |
| RPS6KA3 | 2S(SC+S) ; Definitive | set-1 | . |
| RSRC1 | S; . | set-1 | . |
| SATB1 | 1S(HC+S) ; Definitive | set-1 | . |
| SATB2 | 2S(SC+S) ; Definitive | set-1 | . |
| SCAF4 | 2S(SC+S) ; . | set-1 | . |
| SCN1A | 1S(HC+S) ; . | set-1 | . |
| SETD1A | 1S(HC+S) ; . | set-1 | . |
| SETD1B | 2S(SC+S) ; Definitive | set-1 | . |
| SETD5 | 1S(HC+S) ; Definitive | set-1 | . |
| SGSH | S; . | set-1 | . |
| SHANK3 | 1S(HC+S) ; Definitive | set-1 | . |
| SIK1 | S; . | set-1 | . |
| SIN3A | 1S(HC+S) ; Definitive | set-1 | . |
| SIN3B | 2S(SC+S) ; . | set-1 | . |
| SLC45A1 | S; . | set-1 | . |
| SLC6A1 | 1S(HC+S) ; . | set-1 | . |
| SLC9A1 | 3S(SE+S) ; . | set-1 | . |
| SLC9A6 | 1S(HC+S) ; . | set-1 | . |
| SLITRK2 | S; . | set-1 | . |
| SMARCA2 | 1S(HC+S) ; Definitive | set-1 | . |
| SMARCC2 | 1S(HC+S) ; Definitive | set-1 | . |
| SMC1A | S; Definitive | set-1 | . |
| SMC3 | 2S(SC+S) ; Definitive | set-1 | . |
| SNX14 | S; . | set-1 | . |
| SON | 1S(HC+S) ; Definitive | set-1 | . |
| SOX5 | 1S(HC+S) ; Definitive | set-1 | . |

| | | | |
|---|---|---|---|
| SOX6 | S; Definitive | set-1 | . |
| SPTBN1 | 2S(SC+S) ; . | set-1 | . |
| SRRM2 | 2S(SC+S) ; . | set-1 | . |
| SRSF1 | 3S(SE+S) ; . | set-1 | . |
| STAG1 | S; Definitive | set-1 | . |
| STXBP1 | 1S(HC+S) ; . | set-1 | . |
| SUPT16H | 2S(SC+S) ; . | set-1 | . |
| SYNGAP1 | 1S(HC+S) ; . | set-1 | . |
| SYT1 | S; . | set-1 | . |
| TAF1 | S; . | set-1 | . |
| TAF4 | 2S(SC+S) ; . | set-1 | . |
| TANC2 | 1S(HC+S) ; Definitive | set-1 | . |
| TAOK1 | 1S(HC+S) ; Definitive | set-1 | . |
| TBC1D23 | S; . | set-1 | . |
| TBCK | 1S(HC+S) ; Definitive | set-1 | . |
| TBX1 | S; . | set-1 | . |
| TCEAL1 | 3S(SE+S) ; . | set-1 | . |
| TCF20 | 1S(HC+S) ; . | set-1 | . |
| TCF4 | 1S(HC+S) ; . | set-1 | . |
| TET3 | S; . | set-1 | . |
| TFE3 | S; Definitive | set-1 | . |
| TLK2 | 1S(HC+S) ; . | set-1 | . |
| TM4SF20 | S; . | set-1 | . |
| TRAF7 | 1S(HC+S) ; Definitive | set-1 | . |
| TRAPPC6B | S; . | set-1 | . |
| TRIM8 | 3S(SE+S) ; Definitive | set-1 | . |
| TRIP12 | 1S(HC+S) ; Definitive | set-1 | . |
| TRPM3 | S; Definitive | set-1 | . |
| TRRAP | 2S(SC+S) ; Definitive | set-1 | . |
| TSC1 | 1S(HC+S) ; . | set-1 | . |
| TSC2 | 1S(HC+S) ; . | set-1 | . |

| | | | |
|---|---|---|---|
| TTI2 | S; . | set-1 | . |
| TTN | 2S(SC+S) ; . | set-1 | . |
| UBE3A | 1S(HC+S) ; . | set-1 | . |
| UNC13A | 2S(SC+S) ; . | set-1 | . |
| UPF3B | 1S(HC+S) ; Definitive | set-1 | . |
| USP7 | 2S(SC+S) ; . | set-1 | . |
| USP9X | 1S(HC+S) ; Definitive | set-1 | . |
| VAMP2 | S; . | set-1 | . |
| VPS13B | 1S(HC+S) ; Definitive | set-1 | . |
| WAC | 1S(HC+S) ; Definitive | set-1 | . |
| WASF1 | S; . | set-1 | . |
| WDR26 | S; Definitive | set-1 | . |
| WDR5 | S; . | set-1 | . |
| XPC | S; . | set-1 | . |
| YWHAG | 3S(SE+S) ; . | set-1 | . |
| YY1 | 1S(HC+S) ; . | set-1 | . |
| ZBTB18 | S; Definitive | set-1 | . |
| ZBTB20 | 1S(HC+S) ; Definitive | set-1 | . |
| ZBTB7A | S; . | set-1 | . |
| ZFHX3 | 3S(SE+S) ; . | set-1 | . |
| ZFX | 3S(SE+S) ; Strong | set-1 | . |
| ZMIZ1 | 2S(SC+S) ; Definitive | set-1 | . |
| ZMYM2 | 2S(SC+S) ; . | set-1 | . |
| ZMYM3 | S; . | set-1 | . |
| ZMYND8 | 1S(HC+S) ; . | set-1 | . |
| ZNF292 | 1S(HC+S) ; Definitive | set-1 | . |
| ZNF462 | 1S(HC+S) ; . | set-1 | . |
| ZSWIM6 | S; . | set-1 | . |
| ABCE1 | HC; . | set-1 | . |
| AFF2 | HC; Definitive | set-1 | . |
| ANK2 | HC; Definitive | set-1 | . |

| | | | |
|---|---|---|---|
| ANK3 | HC; Moderate | set-1 | . |
| ANP32A | HC; . | set-1 | . |
| AP2S1 | HC; . | set-1 | . |
| ARF3 | HC; . | set-1 | . |
| ASH1L | HC; Definitive | set-1 | . |
| ATRX | HC; Definitive | set-1 | . |
| AUTS2 | HC; Definitive | set-1 | . |
| BAZ2B | HC; Limited | set-1 | . |
| BCKDK | HC; . | set-1 | . |
| CACNA1E | HC; Definitive | set-1 | . |
| CACNA2D3 | HC; . | set-1 | . |
| CAMTA2 | HC; . | set-1 | . |
| CAPRIN1 | HC; . | set-1 | . |
| CASK | HC; Definitive | set-1 | . |
| CASZ1 | HC; . | set-1 | . |
| CELF4 | HC; . | set-1 | . |
| CIC | HC; Definitive | set-1 | . |
| CORO1A | HC; . | set-1 | . |
| CPSF7 | HC; . | set-1 | . |
| CTNNB1 | HC; Definitive | set-1 | . |
| CUL3 | HC; Definitive | set-1 | . |
| DIP2A | HC; . | set-1 | . |
| DLG4 | HC; Definitive | set-1 | . |
| DPYSL2 | HC; . | set-1 | . |
| DSCAM | HC; . | set-1 | . |
| DYNC1H1 | HC; . | set-1 | . |
| EIF3G | HC; . | set-1 | . |
| ELAVL3 | HC; . | set-1 | . |
| FOXP2 | HC; Definitive | set-1 | . |
| GABRB2 | HC; Definitive | set-1 | . |
| GABRB3 | HC; . | set-1 | . |

| | | | |
|---|---|---|---|
| GFAP | HC; . | set-1 | . |
| GIGYF1 | HC; . | set-1 | . |
| GIGYF2 | HC; . | set-1 | . |
| GRIA2 | HC; . | set-1 | . |
| GRIN1 | HC; . | set-1 | . |
| GRIN2A | HC; . | set-1 | . |
| GRIN2B | HC; . | set-1 | . |
| HDLBP | HC; . | set-1 | . |
| HECTD4 | HC; . | set-1 | . |
| HNRNPH2 | HC; Definitive | set-1 | . |
| HRAS | HC; . | set-1 | . |
| KATNAL2 | HC; Disputed | set-1 | . |
| KCNQ3 | HC; . | set-1 | . |
| KDM2B | HC; . | set-1 | . |
| KDM5B | HC; Moderate | set-1 | . |
| KDM5C | HC; Definitive | set-1 | . |
| KDM6B | HC; Limited | set-1 | . |
| KIAA0232 | HC; . | set-1 | . |
| KLHL20 | HC; . | set-1 | . |
| KMT5B | HC; . | set-1 | . |
| LDB1 | HC; . | set-1 | . |
| LRRC4C | HC; . | set-1 | . |
| LZTR1 | HC; . | set-1 | . |
| MAGEC3 | HC; . | set-1 | . |
| MAP1A | HC; . | set-1 | . |
| MKX | HC; . | set-1 | . |
| MYCBP2 | HC; . | set-1 | . |
| MYT1L | HC; Definitive | set-1 | . |
| NCKAP1 | HC; Definitive | set-1 | . |
| NCOA1 | HC; . | set-1 | . |
| NEXMIF | HC; Definitive | set-1 | . |

| | | | |
|---|---|---|---|
| NLGN2 | HC; . | set-1 | . |
| NLGN3 | HC; Moderate | set-1 | . |
| NLGN4X | HC; Definitive | set-1 | . |
| NR4A2 | HC; Definitive | set-1 | . |
| NRXN1 | HC; Definitive | set-1 | . |
| NRXN2 | HC; . | set-1 | . |
| NRXN3 | HC; . | set-1 | . |
| NUP155 | HC; . | set-1 | . |
| PAH | HC; . | set-1 | . |
| PAX5 | HC; . | set-1 | . |
| PHF12 | HC; . | set-1 | . |
| PHF2 | HC; . | set-1 | . |
| PHF3 | HC; . | set-1 | . |
| PPP1R9B | HC; . | set-1 | . |
| PPP5C | HC; . | set-1 | . |
| PRR14L | HC; . | set-1 | . |
| PSMD11 | HC; . | set-1 | . |
| PSMD6 | HC; . | set-1 | . |
| PTCHD1 | HC; Definitive | set-1 | . |
| PTK7 | HC; . | set-1 | . |
| RALGAPB | HC; . | set-1 | . |
| RELN | HC; Disputed | set-1 | . |
| RFX3 | HC; Definitive | set-1 | . |
| RIMS1 | HC; . | set-1 | . |
| RUNX1T1 | HC; . | set-1 | . |
| SCN2A | HC; . | set-1 | . |
| SCN8A | HC; . | set-1 | . |
| SETBP1 | HC; Definitive | set-1 | . |
| SETD2 | HC; Strong | set-1 | . |
| SHANK2 | HC; Definitive | set-1 | . |
| SKI | HC; . | set-1 | . |

| | | | |
|---|---|---|---|
| SMARCA4 | HC; Definitive | set-1 | . |
| SOS2 | HC; . | set-1 | . |
| SPAST | HC; . | set-1 | . |
| SRCAP | HC; Definitive | set-1 | . |
| SRPRA | HC; . | set-1 | . |
| SYN1 | HC; Definitive | set-1 | . |
| TBCEL | HC; . | set-1 | . |
| TBL1XR1 | HC; Definitive | set-1 | . |
| TBR1 | HC; Definitive | set-1 | . |
| TCF7L2 | HC; Definitive | set-1 | . |
| TEK | HC; . | set-1 | . |
| TLE3 | HC; . | set-1 | . |
| TM9SF4 | HC; . | set-1 | . |
| TRIM23 | HC; . | set-1 | . |
| TRIO | HC; Definitive | set-1 | . |
| TSHZ1 | HC; . | set-1 | . |
| TSHZ3 | HC; . | set-1 | . |
| UBAP2L | HC; . | set-1 | . |
| UBR1 | HC; . | set-1 | . |
| VEZF1 | HC; . | set-1 | . |
| WDFY3 | HC; Definitive | set-1 | . |
| ZBTB21 | HC; . | set-1 | . |
| ACSL4 | ..; Definitive | set-1 | . |
| ALG1 | ..; Definitive | set-1 | . |
| ALG12 | ..; Definitive | set-1 | . |
| ALG3 | ..; Definitive | set-1 | . |
| AP1G1 | ..; Strong | set-1 | . |
| AP4B1 | ..; Definitive | set-1 | . |
| AP4E1 | ..; Definitive | set-1 | . |
| AP4M1 | ..; Definitive | set-1 | . |
| AP4S1 | ..; Definitive | set-1 | . |

| | | | |
|---|---|---|---|
| ASXL1 | ..; Definitive | set-1 | . |
| ASXL2 | ..; Definitive | set-1 | . |
| ATP13A2 | ..; Definitive | set-1 | . |
| ATP6AP2 | ..; Definitive | set-1 | . |
| ATP7A | ..; Definitive | set-1 | . |
| BCAP31 | ..; Definitive | set-1 | . |
| BPTF | ..; Definitive | set-1 | . |
| CAMTA1 | ..; Definitive | set-1 | . |
| CRADD | ..; Definitive | set-1 | . |
| DKC1 | ..; Definitive | set-1 | . |
| DLG3 | ..; Definitive | set-1 | . |
| DPF2 | ..; Definitive | set-1 | . |
| EBP | ..; Definitive | set-1 | . |
| EFTUD2 | ..; Definitive | set-1 | . |
| EIF2S3 | ..; Definitive | set-1 | . |
| EZH2 | ..; Definitive | set-1 | . |
| FGD1 | ..; Definitive | set-1 | . |
| FGF12 | ..; Definitive | set-1 | . |
| FOLR1 | ..; Definitive | set-1 | . |
| FTSJ1 | ..; Definitive | set-1 | . |
| GATAD2B | ..; Definitive | set-1 | . |
| GNB1 | ..; Definitive | set-1 | . |
| GPC3 | ..; Definitive | set-1 | . |
| HPRT1 | ..; Definitive | set-1 | . |
| IDS | ..; Definitive | set-1 | . |
| L1CAM | ..; Definitive | set-1 | . |
| LINS1 | ..; Definitive | set-1 | . |
| MAN1B1 | ..; Definitive | set-1 | . |
| MBTPS2 | ..; Definitive | set-1 | . |
| MED12 | ..; Definitive | set-1 | . |
| MID1 | ..; Definitive | set-1 | . |

| | | | |
|---|---|---|---|
| NDP | ..; Definitive | set-1 | . |
| NHS | ..; Definitive | set-1 | . |
| NONO | ..; Definitive | set-1 | . |
| NSUN2 | ..; Definitive | set-1 | . |
| OTUD6B | ..; Definitive | set-1 | . |
| PAK3 | ..; Definitive | set-1 | . |
| PGAP3 | ..; Definitive | set-1 | . |
| PHF6 | ..; Definitive | set-1 | . |
| PIDD1 | ..; Definitive | set-1 | . |
| PIGL | ..; Definitive | set-1 | . |
| PIGN | ..; Definitive | set-1 | . |
| PLP1 | ..; Definitive | set-1 | . |
| PORCN | ..; Definitive | set-1 | . |
| PPP2R1A | ..; Definitive | set-1 | . |
| PQBP1 | ..; Definitive | set-1 | . |
| SLC16A2 | ..; Definitive | set-1 | . |
| SLC2A1 | ..; Definitive | set-1 | . |
| SMARCB1 | ..; Definitive | set-1 | . |
| SMS | ..; Definitive | set-1 | . |
| TELO2 | ..; Definitive | set-1 | . |
| TUBB3 | ..; Definitive | set-1 | . |
| TUSC3 | ..; Definitive | set-1 | . |
| UBE2A | ..; Definitive | set-1 | . |
| UBTF | ..; Definitive | set-1 | . |
| ZC4H2 | ..; Definitive | set-1 | . |
| ZDHHC9 | ..; Definitive | set-1 | . |
| ZEB2 | ..; Definitive | set-1 | . |
| ADNP | . | set-2 | 41.5; Definitive |
| AHDC1 | . | set-2 | 14.25; Definitive |
| ANKRD11 | . | set-2 | 22.6; Definitive |
| ARHGEF9 | . | set-2 | 14.2; Definitive |

| | | | |
|---|---|---|---|
| ARID1B | . | set-2 | 34.75; Definitive |
| ARX | . | set-2 | 13.8; Definitive |
| ASH1L | . | set-2 | 14.15; Definitive |
| ASXL3 | . | set-2 | 28.85; Definitive |
| AUTS2 | . | set-2 | 35.5; Definitive |
| BRAF | . | set-2 | 13.05; . |
| CACNA1D | . | set-2 | 12.7; . |
| CHD2 | . | set-2 | 25; . |
| CHD8 | . | set-2 | 97.65; Definitive |
| CIC | . | set-2 | 18.7; Definitive |
| CNTN6 | . | set-2 | 13.8; Disputed |
| CREBBP | . | set-2 | 31.35; Definitive |
| CSDE1 | . | set-2 | 15.55; Definitive |
| CTNNB1 | . | set-2 | 32.75; Definitive |
| CTTNBP2 | . | set-2 | 24.8; . |
| CUL3 | . | set-2 | 18.4; Definitive |
| DDX3X | . | set-2 | 78.6; Definitive |
| DEAF1 | . | set-2 | 30.9; . |
| DIP2A | . | set-2 | 18.2; . |
| DMD | . | set-2 | 40.95; . |
| DNMT3A | . | set-2 | 15.9; Limited |
| DSCAM | . | set-2 | 13.5; . |
| DYNC1H1 | . | set-2 | 13.45; . |
| DYRK1A | . | set-2 | 20.2; Definitive |
| EHMT1 | . | set-2 | 13.5; Definitive |
| EP300 | . | set-2 | 23.6; . |
| FOXP1 | . | set-2 | 60.45; Definitive |
| GIGYF1 | . | set-2 | 17.25; . |
| GRIN2B | . | set-2 | 29.65; . |
| HNRNPU | . | set-2 | 38.8; Definitive |
| KDM6B | . | set-2 | 13.75; Limited |

| | | | |
|---|---|---|---|
| MARK2 | . | set-2 | 20.05; . |
| MBD5 | . | set-2 | 46.6; Definitive |
| MECP2 | . | set-2 | 106.65; . |
| MED13L | . | set-2 | 35; Definitive |
| MSL3 | . | set-2 | 13.9; Definitive |
| MYT1L | . | set-2 | 20.35; Definitive |
| NAA15 | . | set-2 | 31.7; Definitive |
| NCKAP1 | . | set-2 | 24.75; Definitive |
| NIPBL | . | set-2 | 15.3; Definitive |
| NRXN1 | . | set-2 | 143.75; Definitive |
| NSD1 | . | set-2 | 22.9; Definitive |
| POGZ | . | set-2 | 35.4; Definitive |
| PTCHD1-AS | . | set-2 | 13.1; . |
| PTEN | . | set-2 | 63.15; . |
| RAI1 | . | set-2 | 12.25; Definitive |
| RFX3 | . | set-2 | 15.95; Definitive |
| SATB2 | . | set-2 | 24.95; Definitive |
| SCN2A | . | set-2 | 109.3; . |
| SETD5 | . | set-2 | 28.05; Definitive |
| SHANK1 | . | set-2 | 14.65; Definitive |
| SHANK2 | . | set-2 | 18.55; Definitive |
| SHANK3 | . | set-2 | 74.85; Definitive |
| SLC6A1 | . | set-2 | 31.15; . |
| SOX5 | . | set-2 | 17.6; Definitive |
| STXBP1 | . | set-2 | 19.1; . |
| SYNGAP1 | . | set-2 | 40.75; . |
| TANC2 | . | set-2 | 24.95; Definitive |
| TCF20 | . | set-2 | 38; . |
| TCF4 | . | set-2 | 13.5; . |
| TLK2 | . | set-2 | 13.5; . |
| TRIP12 | . | set-2 | 27; Definitive |

| | | | |
|---|---|---|---|
| TSC1 | . | set-2 | 17.1; . |
| UBR5 | . | set-2 | 18.45; . |
| WAC | . | set-2 | 15; Definitive |
| WDFY3 | . | set-2 | 17.2; Definitive |
| ACSL4 | . | set-2 | .; Definitive |
| ALG1 | . | set-2 | .; Definitive |
| ALG12 | . | set-2 | .; Definitive |
| ALG3 | . | set-2 | .; Definitive |
| ALG6 | . | set-2 | .; Definitive |
| AP1G1 | . | set-2 | .; Strong |
| AP1S2 | . | set-2 | .; Definitive |
| AP4B1 | . | set-2 | .; Definitive |
| AP4E1 | . | set-2 | .; Definitive |
| AP4M1 | . | set-2 | .; Definitive |
| AP4S1 | . | set-2 | .; Definitive |
| ARID1A | . | set-2 | .; Definitive |
| ARID2 | . | set-2 | .; Definitive |
| ASXL1 | . | set-2 | .; Definitive |
| ASXL2 | . | set-2 | .; Definitive |
| ATP13A2 | . | set-2 | .; Definitive |
| ATP6AP2 | . | set-2 | .; Definitive |
| ATP7A | . | set-2 | .; Definitive |
| BCAP31 | . | set-2 | .; Definitive |
| BPTF | . | set-2 | .; Definitive |
| BRD4 | . | set-2 | .; Definitive |
| BRWD3 | . | set-2 | .; Definitive |
| CAMTA1 | . | set-2 | .; Definitive |
| CASK | . | set-2 | .; Definitive |
| CC2D1A | . | set-2 | .; Definitive |
| CDK13 | . | set-2 | .; Definitive |
| CHAMP1 | . | set-2 | .; Definitive |

| | | | |
|---|---|---|---|
| CHD3 | . | set-2 | .; Definitive |
| CHD4 | . | set-2 | .; Definitive |
| CLCN4 | . | set-2 | .; Definitive |
| CNKSR2 | . | set-2 | .; Definitive |
| CNOT1 | . | set-2 | .; Definitive |
| CRADD | . | set-2 | .; Definitive |
| CUL4B | . | set-2 | .; Definitive |
| DHCR7 | . | set-2 | .; Definitive |
| DHX30 | . | set-2 | .; Definitive |
| DKC1 | . | set-2 | .; Definitive |
| DLG3 | . | set-2 | .; Definitive |
| DPF2 | . | set-2 | .; Definitive |
| EBP | . | set-2 | .; Definitive |
| EFTUD2 | . | set-2 | .; Definitive |
| EIF2S3 | . | set-2 | .; Definitive |
| EIF3F | . | set-2 | .; Definitive |
| EZH2 | . | set-2 | .; Definitive |
| FGD1 | . | set-2 | .; Definitive |
| FGF12 | . | set-2 | .; Definitive |
| FLNA | . | set-2 | .; Definitive |
| FMR1 | . | set-2 | .; Definitive |
| FOLR1 | . | set-2 | .; Definitive |
| FOXP2 | . | set-2 | .; Definitive |
| FRMPD4 | . | set-2 | .; Definitive |
| FTSJ1 | . | set-2 | .; Definitive |
| GATAD2B | . | set-2 | .; Definitive |
| GNB1 | . | set-2 | .; Definitive |
| GPC3 | . | set-2 | .; Definitive |
| GRIA3 | . | set-2 | .; Definitive |
| H1-4 | . | set-2 | .; Definitive |
| HCFC1 | . | set-2 | .; Definitive |

| | | | |
|---|---|---|---|
| HECW2 | . | set-2 | .; Definitive |
| HEPACAM | . | set-2 | .; Definitive |
| HNRNPH2 | . | set-2 | .; Definitive |
| HNRNPK | . | set-2 | .; Definitive |
| HNRNPR | . | set-2 | .; Definitive |
| HOXA1 | . | set-2 | .; Definitive |
| HPRT1 | . | set-2 | .; Definitive |
| HUWE1 | . | set-2 | .; Definitive |
| IDS | . | set-2 | .; Definitive |
| IL1RAPL1 | . | set-2 | .; Definitive |
| IQSEC2 | . | set-2 | .; Definitive |
| ITPR1 | . | set-2 | .; Definitive |
| KAT6A | . | set-2 | .; Definitive |
| KAT6B | . | set-2 | .; Definitive |
| KCNC2 | . | set-2 | .; Definitive |
| KCNH1 | . | set-2 | .; Definitive |
| KDM5C | . | set-2 | .; Definitive |
| KIF1A | . | set-2 | .; Definitive |
| KMT2B | . | set-2 | .; Definitive |
| KMT2C | . | set-2 | .; Definitive |
| KMT2E | . | set-2 | .; Definitive |
| L1CAM | . | set-2 | .; Definitive |
| LINS1 | . | set-2 | .; Definitive |
| MAGEL2 | . | set-2 | .; Definitive |
| MAN1B1 | . | set-2 | .; Definitive |
| MAOA | . | set-2 | .; Definitive |
| MBTPS2 | . | set-2 | .; Definitive |
| MCPH1 | . | set-2 | .; Definitive |
| MED12 | . | set-2 | .; Definitive |
| MID1 | . | set-2 | .; Definitive |
| NAA10 | . | set-2 | .; Definitive |

| | | | |
|---|---|---|---|
| NACC1 | . | set-2 | .; Definitive |
| NBEA | . | set-2 | .; Definitive |
| NDP | . | set-2 | .; Definitive |
| NEXMIF | . | set-2 | .; Definitive |
| NFIB | . | set-2 | .; Definitive |
| NHS | . | set-2 | .; Definitive |
| NONO | . | set-2 | .; Definitive |
| NR2F1 | . | set-2 | .; Definitive |
| NSD2 | . | set-2 | .; Definitive |
| NSUN2 | . | set-2 | .; Definitive |
| OCRL | . | set-2 | .; Definitive |
| OPHN1 | . | set-2 | .; Definitive |
| OTUD6B | . | set-2 | .; Definitive |
| PACS2 | . | set-2 | .; Definitive |
| PAK3 | . | set-2 | .; Definitive |
| PGAP3 | . | set-2 | .; Definitive |
| PHF6 | . | set-2 | .; Definitive |
| PHF8 | . | set-2 | .; Definitive |
| PHIP | . | set-2 | .; Definitive |
| PIDD1 | . | set-2 | .; Definitive |
| PIGL | . | set-2 | .; Definitive |
| PIGN | . | set-2 | .; Definitive |
| PLP1 | . | set-2 | .; Definitive |
| PORCN | . | set-2 | .; Definitive |
| POU3F3 | . | set-2 | .; Definitive |
| PPM1D | . | set-2 | .; Definitive |
| PPP2R1A | . | set-2 | .; Definitive |
| PQBP1 | . | set-2 | .; Definitive |
| PUF60 | . | set-2 | .; Definitive |
| QRICH1 | . | set-2 | .; Definitive |
| RAB39B | . | set-2 | .; Definitive |

| | | | |
|---|---|---|---|
| RAC1 | . | set-2 | .; Definitive |
| RAD21 | . | set-2 | .; Definitive |
| RALA | . | set-2 | .; Definitive |
| RFX7 | . | set-2 | .; Definitive |
| RPL10 | . | set-2 | .; Definitive |
| RPS6KA3 | . | set-2 | .; Definitive |
| SETBP1 | . | set-2 | .; Definitive |
| SETD1B | . | set-2 | .; Definitive |
| SETD2 | . | set-2 | .; Strong |
| SIN3A | . | set-2 | .; Definitive |
| SLC16A2 | . | set-2 | .; Definitive |
| SLC2A1 | . | set-2 | .; Definitive |
| SMAD4 | . | set-2 | .; Definitive |
| SMARCA2 | . | set-2 | .; Definitive |
| SMARCA4 | . | set-2 | .; Definitive |
| SMARCB1 | . | set-2 | .; Definitive |
| SMC1A | . | set-2 | .; Definitive |
| SMC3 | . | set-2 | .; Definitive |
| SMS | . | set-2 | .; Definitive |
| SNAP25 | . | set-2 | .; Definitive |
| SOX6 | . | set-2 | .; Definitive |
| SRCAP | . | set-2 | .; Definitive |
| STAG1 | . | set-2 | .; Definitive |
| TELO2 | . | set-2 | .; Definitive |
| TFE3 | . | set-2 | .; Definitive |
| TNRC6B | . | set-2 | .; Definitive |
| TRAPPC9 | . | set-2 | .; Definitive |
| TRIM8 | . | set-2 | .; Definitive |
| TRIO | . | set-2 | .; Definitive |
| TRPM3 | . | set-2 | .; Definitive |
| TRRAP | . | set-2 | .; Definitive |

| | | | |
|---|---|---|---|
| TUBB3 | . | set-2 | .; Definitive |
| TUSC3 | . | set-2 | .; Definitive |
| UBE2A | . | set-2 | .; Definitive |
| UBTF | . | set-2 | .; Definitive |
| UPF3B | . | set-2 | .; Definitive |
| USP9X | . | set-2 | .; Definitive |
| WDR26 | . | set-2 | .; Definitive |
| ZBTB18 | . | set-2 | .; Definitive |
| ZBTB20 | . | set-2 | .; Definitive |
| ZC4H2 | . | set-2 | .; Definitive |
| ZDHHC9 | . | set-2 | .; Definitive |
| ZEB2 | . | set-2 | .; Definitive |
| ZFX | . | set-2 | .; Strong |
| ZMIZ1 | . | set-2 | .; Definitive |
| ZMYND11 | . | set-2 | .; Definitive |
| ZNF292 | . | set-2 | .; Definitive |

| **Supplementary Table 2:** Predicted damaging genetic variants identified from the IndGenomes dataset using the ASD gene panel (517 genes), filtered using REVEL scores >0.75 for missense variants and LoFTEE-predicted high-confidence loss-of-function (LoF) variants. | | | |
|---|---|---|---|
| **Variant** | **Gene** | **Class** | **Prediction** |
| chr1-8335564-G-A | SLC45A1 | nonsynonymous | 0.803 |
| chr1-8339522-A-T | SLC45A1 | nonsynonymous | 0.85 |
| chr1-16988409-C-T | ATP13A2 | nonsynonymous | 0.907 |
| chr1-16996058-C-T | ATP13A2 | nonsynonymous | 0.916 |
| chr1-16997104-G-A | ATP13A2 | nonsynonymous | 0.779 |
| chr1-17004366-A-C | ATP13A2 | nonsynonymous | 0.816 |
| chr1-39282355-G-A | MACF1 | nonsynonymous | 0.881 |
| chr1-39282356-C-T | MACF1 | nonsynonymous | 0.918 |
| chr1-39283299-A-T | MACF1 | nonsynonymous | 0.919 |
| chr1-39287321-G-A | MACF1 | nonsynonymous | 0.766 |
| chr1-39287339-T-C | MACF1 | nonsynonymous | 0.929 |
| chr1-39293533-T-G | MACF1 | nonsynonymous | 0.844 |
| chr1-42930631-C-T | SLC2A1 | nonsynonymous | 0.832 |
| chr1-46192183-C-T | POMGNT1 | nonsynonymous | 0.818 |
| chr1-63396585-C-T | ALG6 | nonsynonymous | 0.892 |
| chr1-63402322-A-C | ALG6 | nonsynonymous | 0.87 |
| chr1-63406340-T-G | ALG6 | nonsynonymous | 0.834 |
| chr1-63415884-C-T | ALG6 | nonsynonymous | 0.912 |
| chr1-116390252-A-T | ATP1A1 | nonsynonymous | 0.933 |
| chr1-116390255-T-A | ATP1A1 | nonsynonymous | 0.938 |
| chr1-181483860-A-G | CACNA1E | nonsynonymous | 0.753 |
| chr1-181580763-C-T | CACNA1E | nonsynonymous | 0.755 |
| chr1-181739175-A-G | CACNA1E | nonsynonymous | 0.851 |
| chr1-181758828-C-T | CACNA1E | nonsynonymous | 0.924 |
| chr1-202760439-C-G | KDM5B | nonsynonymous | 0.876 |
| chr1-224411554-G-A | WDR26 | nonsynonymous | 0.788 |
| chr1-244858234-T-C | HNRNPU | nonsynonymous | 0.82 |
| chr10-74981898-T-G | KAT6B | nonsynonymous | 0.813 |

| | | | |
|---|---|---|---|
| chr10-77980183-G-A | POLR3A | nonsynonymous | 0.826 |
| chr10-77984287-A-G | POLR3A | nonsynonymous | 0.816 |
| chr10-77985960-C-T | POLR3A | nonsynonymous | 0.753 |
| chr10-78001031-C-T | POLR3A | nonsynonymous | 0.801 |
| chr10-113143932-G-T | TCF7L2 | nonsynonymous | 0.924 |
| chr11-799858-C-T | PIDD1 | nonsynonymous | 0.826 |
| chr11-7995318-C-G | EIF3F | nonsynonymous | 0.776 |
| chr11-61420482-C-T | CPSF7 | nonsynonymous | 0.763 |
| chr11-63899921-C-A | MARK2 | nonsynonymous | 0.911 |
| chr11-64660377-G-A | NRXN2 | nonsynonymous | 0.816 |
| chr11-71435422-G-A | DHCR7 | nonsynonymous | 0.772 |
| chr11-71435454-C-T | DHCR7 | nonsynonymous | 0.793 |
| chr11-71438967-C-A | DHCR7 | nonsynonymous | 0.851 |
| chr11-71439030-G-A | DHCR7 | nonsynonymous | 0.988 |
| chr11-71441416-T-C | DHCR7 | nonsynonymous | 0.822 |
| chr11-71442319-T-C | DHCR7 | nonsynonymous | 0.858 |
| chr11-71444162-G-A | DHCR7 | nonsynonymous | 0.958 |
| chr11-118504444-A-C | KMT2A | nonsynonymous | 0.751 |
| chr11-118506180-T-G | KMT2A | nonsynonymous | 0.822 |
| chr12-2593255-G-A | CACNA1C | nonsynonymous | 0.779 |
| chr12-2610673-C-T | CACNA1C | nonsynonymous | 0.878 |
| chr12-2674633-C-T | CACNA1C | nonsynonymous | 0.784 |
| chr12-6570907-G-A | CHD4 | nonsynonymous | 0.935 |
| chr12-6577877-G-A | CHD4 | nonsynonymous | 0.753 |
| chr12-6583302-C-A | CHD4 | nonsynonymous | 0.814 |
| chr12-6601715-C-G | CHD4 | nonsynonymous | 0.822 |
| chr12-13753704-G-A | GRIN2B | nonsynonymous | 0.782 |
| chr12-23741026-G-T | SOX5 | nonsynonymous | 0.776 |
| chr12-51687096-C-T | SCN8A | nonsynonymous | 0.918 |
| chr12-51769121-T-G | SCN8A | nonsynonymous | 0.891 |
| chr12-75051073-A-G | KCNC2 | nonsynonymous | 0.76 |

| | | | |
|---|---|---|---|
| chr12-89620194-G-A | ATP2B1 | nonsynonymous | 0.807 |
| chr12-89655798-G-A | ATP2B1 | nonsynonymous | 0.759 |
| chr12-102844410-A-G | PAH | nonsynonymous | 0.983 |
| chr12-102855285-G-A | PAH | nonsynonymous | 0.87 |
| chr12-102877548-G-A | PAH | nonsynonymous | 0.954 |
| chr12-102912801-C-T | PAH | nonsynonymous | 0.789 |
| chr12-112486532-G-A | PTPN11 | nonsynonymous | 0.906 |
| chr12-116019400-A-G | MED13L | nonsynonymous | 0.86 |
| chr13-35566908-G-A | NBEA | nonsynonymous | 0.865 |
| chr13-35646296-C-T | NBEA | nonsynonymous | 0.937 |
| chr13-77158076-C-T | MYCBP2 | nonsynonymous | 0.763 |
| chr13-100257606-A-G | PCCA | nonsynonymous | 0.94 |
| chr13-100302927-C-G | PCCA | nonsynonymous | 0.755 |
| chr13-100330561-G-A | PCCA | nonsynonymous | 0.835 |
| chr14-21366508-T-C | SUPT16H | nonsynonymous | 0.847 |
| chr14-21369811-G-A | SUPT16H | nonsynonymous | 0.856 |
| chr14-29597634-A-C | PRKD1 | nonsynonymous | 0.922 |
| chr14-29597706-C-T | PRKD1 | nonsynonymous | 0.755 |
| chr14-29630927-G-A | PRKD1 | nonsynonymous | 0.798 |
| chr14-29666098-G-A | PRKD1 | nonsynonymous | 0.823 |
| chr14-29666172-T-C | PRKD1 | nonsynonymous | 0.968 |
| chr14-50160018-T-C | SOS2 | nonsynonymous | 0.82 |
| chr14-50201056-G-A | SOS2 | nonsynonymous | 0.825 |
| chr14-78243217-C-T | NRXN3 | nonsynonymous | 0.815 |
| chr14-78709241-C-T | NRXN3 | nonsynonymous | 0.781 |
| chr14-78709422-G-A | NRXN3 | nonsynonymous | 0.852 |
| chr15-28135607-A-G | HERC2 | nonsynonymous | 0.862 |
| chr15-43036228-T-C | UBR1 | nonsynonymous | 0.879 |
| chr15-63632727-C-T | HERC1 | nonsynonymous | 0.779 |
| chr15-63755316-A-C | HERC1 | nonsynonymous | 0.77 |
| chr15-92946160-A-T | CHD2 | nonsynonymous | 0.807 |

| | | | |
|---|---|---|---|
| chr15-92991492-A-G | CHD2 | nonsynonymous | 0.777 |
| chr15-92991495-A-C | CHD2 | nonsynonymous | 0.785 |
| chr16-1495500-C-T | TELO2 | nonsynonymous | 0.804 |
| chr16-2054402-A-G | TSC2 | nonsynonymous | 0.797 |
| chr16-2058770-T-G | TSC2 | nonsynonymous | 0.867 |
| chr16-2070474-C-G | TSC2 | nonsynonymous | 0.928 |
| chr16-2071535-G-A | TSC2 | nonsynonymous | 0.895 |
| chr16-2072361-A-G | TSC2 | nonsynonymous | 0.825 |
| chr16-2079290-A-G | TSC2 | nonsynonymous | 0.924 |
| chr16-2085004-A-T | TSC2 | nonsynonymous | 0.831 |
| chr16-2085263-G-A | TSC2 | nonsynonymous | 0.933 |
| chr16-2086797-C-T | TSC2 | nonsynonymous | 0.951 |
| chr16-2088563-C-T | TSC2 | nonsynonymous | 0.756 |
| chr16-2088564-G-A | TSC2 | nonsynonymous | 0.775 |
| chr16-2088569-C-T | TSC2 | nonsynonymous | 0.812 |
| chr16-3740427-C-T | CREBBP | nonsynonymous | 0.841 |
| chr16-5081056-G-C | ALG1 | nonsynonymous | 0.88 |
| chr16-30185243-C-T | CORO1A | nonsynonymous | 0.905 |
| chr16-30966990-G-T | SETD1A | nonsynonymous | 0.775 |
| chr16-72794668-C-A | ZFHX3 | nonsynonymous | 0.915 |
| chr16-89288565-G-A | ANKRD11 | nonsynonymous | 0.888 |
| chr16-89935409-C-T | TUBB3 | nonsynonymous | 0.794 |
| chr17-1659483-T-C | PRPF8 | nonsynonymous | 0.905 |
| chr17-1677596-C-A | PRPF8 | nonsynonymous | 0.891 |
| chr17-7417312-C-T | NLGN2 | nonsynonymous | 0.795 |
| chr17-7899056-G-A | CHD3 | nonsynonymous | 0.819 |
| chr17-7908504-T-G | CHD3 | nonsynonymous | 0.949 |
| chr17-16316686-T-C | PIGL | nonsynonymous | 0.791 |
| chr17-31337406-T-C | NF1 | nonsynonymous | 0.854 |
| chr17-39673142-C-T | PGAP3 | nonsynonymous | 0.766 |
| chr17-44913428-C-A | GFAP | nonsynonymous | 0.802 |

| | | | |
|---|---|---|---|
| chr17-44915081-G-C | GFAP | nonsynonymous | 0.873 |
| chr17-44915243-T-A | GFAP | nonsynonymous | 0.866 |
| chr17-61960925-T-C | MED13 | nonsynonymous | 0.958 |
| chr17-61961069-A-G | MED13 | nonsynonymous | 0.927 |
| chr17-61962840-G-A | MED13 | nonsynonymous | 0.807 |
| chr17-80210721-G-A | SGSH | nonsynonymous | 0.789 |
| chr17-80217121-C-T | SGSH | nonsynonymous | 0.892 |
| chr17-80220247-G-A | SGSH | nonsynonymous | 0.813 |
| chr18-36138298-A-G | ELP2 | nonsynonymous | 0.766 |
| chr18-36156574-C-T | ELP2 | nonsynonymous | 0.806 |
| chr18-44952639-A-G | SETBP1 | nonsynonymous | 0.76 |
| chr18-47069497-T-C | KATNAL2 | nonsynonymous | 0.928 |
| chr18-47069515-C-A | KATNAL2 | nonsynonymous | 0.904 |
| chr18-47069599-G-A | KATNAL2 | nonsynonymous | 0.938 |
| chr18-47077374-G-A | KATNAL2 | nonsynonymous | 0.851 |
| chr18-62147057-T-A | PIGN | nonsynonymous | 0.808 |
| chr18-62154573-A-T | PIGN | nonsynonymous | 0.911 |
| chr18-62157747-G-A | PIGN | nonsynonymous | 0.79 |
| chr18-62161187-G-A | PIGN | nonsynonymous | 0.757 |
| chr19-13207953-C-G | CACNA1A | nonsynonymous | 0.755 |
| chr19-13208857-G-A | CACNA1A | nonsynonymous | 0.761 |
| chr19-13255124-C-T | CACNA1A | nonsynonymous | 0.76 |
| chr19-14151246-G-A | ADGRL1 | nonsynonymous | 0.792 |
| chr19-14162686-C-T | ADGRL1 | nonsynonymous | 0.769 |
| chr19-14162864-C-T | ADGRL1 | nonsynonymous | 0.783 |
| chr19-14163268-C-T | ADGRL1 | nonsynonymous | 0.806 |
| chr19-14163325-C-G | ADGRL1 | nonsynonymous | 0.843 |
| chr19-17648500-G-A | UNC13A | nonsynonymous | 0.795 |
| chr19-31276963-T-C | TSHZ3 | nonsynonymous | 0.849 |
| chr2-25240693-C-T | DNMT3A | nonsynonymous | 0.965 |
| chr2-50497612-C-A | NRXN1 | nonsynonymous | 0.929 |

| | | | |
|---|---|---|---|
| chr2-50531251-G-A | NRXN1 | nonsynonymous | 0.85 |
| chr2-50538558-A-C | NRXN1 | nonsynonymous | 0.863 |
| chr2-50552596-C-T | NRXN1 | nonsynonymous | 0.78 |
| chr2-119915198-T-G | PTPN4 | nonsynonymous | 0.847 |
| chr2-119920130-T-C | PTPN4 | nonsynonymous | 0.954 |
| chr2-119965560-G-C | PTPN4 | nonsynonymous | 0.822 |
| chr2-156326280-C-G | NR4A2 | nonsynonymous | 0.82 |
| chr2-159428381-C-G | BAZ2B | nonsynonymous | 0.802 |
| chr2-165309366-T-C | SCN2A | nonsynonymous | 0.939 |
| chr2-165386759-G-C | SCN2A | nonsynonymous | 0.805 |
| chr2-165991594-A-G | SCN1A | nonsynonymous | 0.89 |
| chr2-165991694-G-A | SCN1A | nonsynonymous | 0.893 |
| chr2-166013784-G-A | SCN1A | nonsynonymous | 0.822 |
| chr2-166037995-C-A | SCN1A | nonsynonymous | 0.953 |
| chr2-166046880-C-A | SCN1A | nonsynonymous | 0.889 |
| chr2-166051949-T-C | SCN1A | nonsynonymous | 0.814 |
| chr2-178531025-C-T | TTN | nonsynonymous | 0.786 |
| chr2-178534433-C-T | TTN | nonsynonymous | 0.799 |
| chr2-178537377-C-T | TTN | nonsynonymous | 0.797 |
| chr2-178541369-A-G | TTN | nonsynonymous | 0.894 |
| chr2-178549635-G-T | TTN | nonsynonymous | 0.753 |
| chr2-178574742-T-C | TTN | nonsynonymous | 0.882 |
| chr2-178578057-C-G | TTN | nonsynonymous | 0.761 |
| chr2-178581603-A-G | TTN | nonsynonymous | 0.916 |
| chr2-178592210-C-G | TTN | nonsynonymous | 0.765 |
| chr2-178599655-A-G | TTN | nonsynonymous | 0.85 |
| chr2-178602350-G-T | TTN | nonsynonymous | 0.777 |
| chr2-178607496-A-G | TTN | nonsynonymous | 0.801 |
| chr2-178611061-C-A | TTN | nonsynonymous | 0.77 |
| chr2-178612346-C-T | TTN | nonsynonymous | 0.795 |
| chr2-178718968-G-A | TTN | nonsynonymous | 0.768 |

| | | | |
|---|---|---|---|
| chr2-178728383-G-T | TTN | nonsynonymous | 0.82 |
| chr2-178733829-A-C | TTN | nonsynonymous | 0.803 |
| chr2-178746087-A-C | TTN | nonsynonymous | 0.882 |
| chr2-178751667-T-C | TTN | nonsynonymous | 0.786 |
| chr2-178764746-G-A | TTN | nonsynonymous | 0.801 |
| chr2-178769873-C-T | TTN | nonsynonymous | 0.786 |
| chr2-218809598-G-A | CYP27A1 | nonsynonymous | 0.85 |
| chr2-218809731-G-A | CYP27A1 | nonsynonymous | 0.767 |
| chr2-218814409-G-A | CYP27A1 | nonsynonymous | 0.89 |
| chr2-224506991-A-G | CUL3 | nonsynonymous | 0.801 |
| chr2-224557750-T-G | CUL3 | nonsynonymous | 0.859 |
| chr2-240788211-G-A | KIF1A | nonsynonymous | 0.757 |
| chr20-38574774-G-A | RALGAPB | nonsynonymous | 0.841 |
| chr22-20995827-G-T | LZTR1 | nonsynonymous | 0.828 |
| chr22-20995968-T-C | LZTR1 | nonsynonymous | 0.943 |
| chr22-40358874-C-A | ADSL | nonsynonymous | 0.76 |
| chr22-40361289-G-A | ADSL | nonsynonymous | 0.926 |
| chr22-40361511-C-T | ADSL | nonsynonymous | 0.76 |
| chr22-40364950-A-G | ADSL | nonsynonymous | 0.842 |
| chr22-40365025-C-A | ADSL | nonsynonymous | 0.927 |
| chr22-41168803-G-C | EP300 | nonsynonymous | 0.852 |
| chr22-41170537-A-G | EP300 | nonsynonymous | 0.801 |
| chr3-1321791-C-G | CNTN6 | nonsynonymous | 0.799 |
| chr3-4645612-C-A | ITPR1 | nonsynonymous | 0.799 |
| chr3-4667450-C-G | ITPR1 | nonsynonymous | 0.783 |
| chr3-4683655-G-T | ITPR1 | nonsynonymous | 0.814 |
| chr3-49044422-A-T | QRICH1 | nonsynonymous | 0.779 |
| chr3-51986492-G-C | ABHD14A-ACY1,ACY1 | nonsynonymous | 0.849 |
| chr3-51988806-G-A | ABHD14A-ACY1,ACY1 | nonsynonymous | 0.79 |
| chr3-51988813-G-A | ABHD14A-ACY1,ACY1 | nonsynonymous | 0.762 |
| chr3-53723449-T-C | CACNA1D | nonsynonymous | 0.831 |

| | | | |
|---|---|---|---|
| chr3-53732893-T-G | CACNA1D | nonsynonymous | 0.867 |
| chr3-53770518-G-T | CACNA1D | nonsynonymous | 0.975 |
| chr3-54123523-T-C | CACNA2D3 | nonsynonymous | 0.776 |
| chr3-136255919-G-A | PCCB | nonsynonymous | 0.851 |
| chr3-136283888-C-T | PCCB | nonsynonymous | 0.913 |
| chr3-136293762-T-G | PCCB | nonsynonymous | 0.897 |
| chr3-136298003-G-A | PCCB | nonsynonymous | 0.955 |
| chr3-136301034-C-T | PCCB | nonsynonymous | 0.908 |
| chr3-136316991-C-G | PCCB | nonsynonymous | 0.885 |
| chr3-136327706-G-A | PCCB | nonsynonymous | 0.96 |
| chr3-136329928-C-A | PCCB | nonsynonymous | 0.921 |
| chr3-151360553-G-T | MED12L | nonsynonymous | 0.797 |
| chr3-184243833-T-C | ALG3 | nonsynonymous | 0.907 |
| chr3-192360469-T-A | FGF12 | nonsynonymous | 0.875 |
| chr4-1975294-G-A | NSD2 | nonsynonymous | 0.906 |
| chr4-113360859-T-G | ANK2 | nonsynonymous | 0.822 |
| chr4-148120286-T-C | NR3C2 | nonsynonymous | 0.871 |
| chr4-157332871-G-T | GRIA2 | nonsynonymous | 0.85 |
| chr4-157335861-G-T | GRIA2 | nonsynonymous | 0.819 |
| chr5-14286912-T-C | TRIO | nonsynonymous | 0.866 |
| chr5-14290825-G-T | TRIO | nonsynonymous | 0.867 |
| chr5-37291980-A-G | NUP155 | nonsynonymous | 0.805 |
| chr5-37310647-A-G | NUP155 | nonsynonymous | 0.805 |
| chr5-37341182-A-G | NUP155 | nonsynonymous | 0.792 |
| chr5-75381182-C-T | CERT1 | nonsynonymous | 0.887 |
| chr5-98873689-C-A | CHD1 | nonsynonymous | 0.752 |
| chr6-24495228-G-C | ALDH5A1 | nonsynonymous | 0.833 |
| chr6-24522774-T-C | ALDH5A1 | nonsynonymous | 0.786 |
| chr6-24522813-A-C | ALDH5A1 | nonsynonymous | 0.864 |
| chr6-24533615-T-G | ALDH5A1 | nonsynonymous | 0.953 |
| chr6-43132636-G-A | PTK7 | nonsynonymous | 0.854 |

| | | | |
|---|---|---|---|
| chr6-43139478-G-C | PTK7 | nonsynonymous | 0.766 |
| chr6-43158904-C-G | PTK7 | nonsynonymous | 0.887 |
| chr6-72400542-T-A | RIMS1 | nonsynonymous | 0.822 |
| chr6-170283037-G-A | DLL1 | nonsynonymous | 0.816 |
| chr6-170285099-C-T | DLL1 | nonsynonymous | 0.976 |
| chr6-170285134-T-C | DLL1 | nonsynonymous | 0.9 |
| chr7-44242340-A-C | CAMK2B | nonsynonymous | 0.781 |
| chr7-99005207-A-G | TRRAP | nonsynonymous | 0.853 |
| chr7-100678449-C-T | GNB2 | nonsynonymous | 0.776 |
| chr7-100683895-G-C | GIGYF1 | nonsynonymous | 0.763 |
| chr7-114659367-A-T | FOXP2 | nonsynonymous | 0.855 |
| chr7-140777080-C-A | BRAF | nonsynonymous | 0.791 |
| chr7-152144736-A-G | KMT2C | nonsynonymous | 0.887 |
| chr7-152187315-G-T | KMT2C | nonsynonymous | 0.778 |
| chr7-152187324-G-T | KMT2C | nonsynonymous | 0.87 |
| chr8-26627251-G-A | DPYSL2 | nonsynonymous | 0.758 |
| chr8-26643481-C-T | DPYSL2 | nonsynonymous | 0.845 |
| chr8-26644048-A-G | DPYSL2 | nonsynonymous | 0.785 |
| chr8-60820032-A-T | CHD7 | nonsynonymous | 0.824 |
| chr8-60830380-T-A | CHD7 | nonsynonymous | 0.956 |
| chr8-60850517-G-A | CHD7 | nonsynonymous | 0.803 |
| chr8-60850535-G-C | CHD7 | nonsynonymous | 0.822 |
| chr8-60852042-G-A | CHD7 | nonsynonymous | 0.774 |
| chr8-60852533-T-C | CHD7 | nonsynonymous | 0.777 |
| chr8-99156724-A-G | VPS13B | nonsynonymous | 0.891 |
| chr8-104251687-C-A | RIMS2 | nonsynonymous | 0.756 |
| chr8-132141177-G-A | KCNQ3 | nonsynonymous | 0.887 |
| chr8-132172600-G-T | KCNQ3 | nonsynonymous | 0.895 |
| chr8-132180228-G-A | KCNQ3 | nonsynonymous | 0.966 |
| chr9-37015022-C-A | PAX5 | nonsynonymous | 0.871 |
| chr9-70603406-G-A | TRPM3 | nonsynonymous | 0.814 |

| | | | |
|---|---|---|---|
| chr9-98371488-G-T | GABBR2 | nonsynonymous | 0.874 |
| chr9-98385644-C-A | GABBR2 | nonsynonymous | 0.949 |
| chr9-128945893-T-G | DOLK | nonsynonymous | 0.853 |
| chr9-128946564-A-G | DOLK | nonsynonymous | 0.876 |
| chr9-137101641-T-G | MAN1B1 | nonsynonymous | 0.959 |
| chrX-10213798-C-G | CLCN4 | nonsynonymous | 0.758 |
| chrX-32573830-C-A | DMD | nonsynonymous | 0.809 |
| chrX-48524069-C-A | EBP | nonsynonymous | 0.832 |
| chrX-48528281-G-A | EBP | nonsynonymous | 0.945 |
| chrX-71381844-G-T | TAF1 | nonsynonymous | 0.785 |
| chrX-77989278-T-G | ATP7A | nonsynonymous | 0.791 |
| chrX-78011247-A-G | ATP7A | nonsynonymous | 0.776 |
| chrX-78015836-C-T | ATP7A | nonsynonymous | 0.757 |
| chrX-78020274-G-A | ATP7A | nonsynonymous | 0.758 |
| chrX-103790561-C-G | PLP1 | nonsynonymous | 0.867 |
| chrX-154357620-G-A | FLNA | nonsynonymous | 0.831 |
| chrX-154359041-C-T | FLNA | nonsynonymous | 0.897 |
| chrX-154361561-A-G | FLNA | nonsynonymous | 0.797 |
| chrX-154362289-T-C | FLNA | nonsynonymous | 0.833 |
| chrX-154362718-C-T | FLNA | nonsynonymous | 0.931 |
| chrX-154366746-C-T | FLNA | nonsynonymous | 0.828 |
| chrX-154768353-T-C | DKC1 | nonsynonymous | 0.803 |
| chr1-7249583-C-CAGCCTCT | CAMTA1 | frameshift insertion | HC LoF variant |
| chr1-7249585-A-AAATACAG | CAMTA1 | frameshift insertion | HC LoF variant |
| chr1-8324503-C-CG | SLC45A1 | frameshift insertion | HC LoF variant |
| chr1-8324504-T-TG | SLC45A1 | frameshift insertion | HC LoF variant |
| chr1-8324521-CAA-C | SLC45A1 | frameshift deletion | HC LoF variant |
| chr1-10639091-CGT-C | CASZ1 | frameshift deletion | HC LoF variant |
| chr1-10639094-CGTCCTCGTC | CASZ1 | frameshift deletion | HC LoF variant |
| chr1-11145046-C-CCTCCAACA | MTOR | splicing | HC LoF variant |
| chr1-16996162-CACCTGGCAC | ATP13A2 | frameshift deletion | HC LoF variant |

| | | | |
|---|---|---|---|
| chr1-17000142-TCTGCAGGCA | ATP13A2 | splicing | HC LoF variant |
| chr1-17005450-C-T | ATP13A2 | stopgain | HC LoF variant |
| chr1-23313700-T-TA | HNRNPR | frameshift insertion | HC LoF variant |
| chr1-23313703-C-CTCCAGAA( | HNRNPR | splicing | HC LoF variant |
| chr1-26775630-G-T | ARID1A | stopgain | HC LoF variant |
| chr1-27101815-G-T | SLC9A1 | stopgain | HC LoF variant |
| chr1-27103255-TC-T | SLC9A1 | frameshift deletion | HC LoF variant |
| chr1-27552023-G-GCC | AHDC1 | frameshift insertion | HC LoF variant |
| chr1-39335135-C-A | MACF1 | stop gain | HC LoF variant |
| chr1-63406401-T-C | ALG6 | splicing | HC LoF variant |
| chr1-107436758-A-AG | NTNG1 | frameshift insertion | HC LoF variant |
| chr1-107436760-G-GTTATAGC | NTNG1 | frameshift insertion | HC LoF variant |
| chr1-113902859-G-T | AP4B1 | stopgain | HC LoF variant |
| chr1-116390250-A-ATTCT | ATP1A1 | frameshift insertion | HC LoF variant |
| chr1-202736393-CTAA-C | KDM5B | splicing | HC LoF variant |
| chr1-202753014-T-TTTGG | KDM5B | frameshift insertion | HC LoF variant |
| chr1-210683292-CA-C | KCNH1 | frameshift deletion | HC LoF variant |
| chr1-224431494-ACGTTT-A | WDR26 | frameshift deletion | HC LoF variant |
| chr1-224433781-C-A | WDR26 | stopgain | HC LoF variant |
| chr1-230236716-C-T | GALNT2 | stopgain | HC LoF variant |
| chr1-244855902-GCCT-G | HNRNPU | frameshift deletion | HC LoF variant |
| chr10-221236-C-A | ZMYND11 | stopgain | HC LoF variant |
| chr10-27735338-T-A | MKX | stopgain | HC LoF variant |
| chr10-60070194-C-A | ANK3 | stopgain | HC LoF variant |
| chr10-60106048-A-AGT | ANK3 | frameshift insertion | HC LoF variant |
| chr10-74970102-G-T | KAT6B | splicing | HC LoF variant |
| chr10-74979257-G-GAGTCTGT | KAT6B | stopgain | HC LoF variant |
| chr10-75028920-GAAGA-G | KAT6B | frameshift deletion | HC LoF variant |
| chr10-75028926-GAAGGGGAA | KAT6B | frameshift deletion | HC LoF variant |
| chr10-75029884-A-AGAAAAG( | KAT6B | frameshift insertion | HC LoF variant |
| chr11-802383-G-GGGTC | PIDD1 | frameshift insertion | HC LoF variant |

| | | | |
|---|---|---|---|
| chr11-72195749-T-C | FOLR1 | splicing | HC LoF variant |
| chr12-6581174-C-T | CHD4 | splicing | HC LoF variant |
| chr12-88049228-ACT-A | CEP290 | frameshift deletion | HC LoF variant |
| chr12-88049337-A-T | CEP290 | stopgain | HC LoF variant |
| chr12-88055666-A-AT | CEP290 | frameshift insertion | HC LoF variant |
| chr12-88058915-C-CTTATATT( | CEP290 | stopgain | HC LoF variant |
| chr12-88058921-G-A | CEP290 | stopgain | HC LoF variant |
| chr12-88058923-A-T | CEP290 | stopgain | HC LoF variant |
| chr12-88068585-G-GTA | CEP290 | frameshift insertion | HC LoF variant |
| chr12-88071890-T-TA | CEP290 | stopgain | HC LoF variant |
| chr12-88077785-T-TTGTGAAT | CEP290 | frameshift insertion | HC LoF variant |
| chr12-88093903-A-AT | CEP290 | frameshift insertion | HC LoF variant |
| chr12-88126425-GA-G | CEP290 | frameshift deletion | HC LoF variant |
| chr12-93678901-G-T | CRADD | stopgain | HC LoF variant |
| chr12-99648404-G-GC | FAM71C | frameshift insertion | HC LoF variant |
| chr12-99648514-ATC-A | FAM71C | frameshift deletion | HC LoF variant |
| chr12-102866665-T-C | PAH | splicing | HC LoF variant |
| chr12-116007637-C-A | MED13L | splicing | HC LoF variant |
| chr12-116015167-G-GTCTAGA | MED13L | frameshift insertion | HC LoF variant |
| chr12-121441068-A-AC | KDM2B | splicing | HC LoF variant |
| chr12-121580827-CTG-C | KDM2B | frameshift deletion | HC LoF variant |
| chr12-121810266-G-T | SETD1B | stopgain | HC LoF variant |
| chr12-121823327-CACCA-C | SETD1B | frameshift deletion | HC LoF variant |
| chr12-121823330-CACCCCTT- | SETD1B | frameshift deletion | HC LoF variant |
| chr13-26337596-A-AAAGTTAT | CDK8 | frameshift insertion | HC LoF variant |
| chr13-26396355-G-GA | CDK8 | splicing | HC LoF variant |
| chr13-35156147-T-TCCTTA | NBEA | frameshift insertion | HC LoF variant |
| chr13-35593385-C-CTTCCTC( | NBEA | frameshift insertion | HC LoF variant |
| chr13-77077265-G-GAT | MYCBP2 | frameshift insertion | HC LoF variant |
| chr13-77146211-C-CT | MYCBP2 | frameshift insertion | HC LoF variant |
| chr13-77326949-TC-T | MYCBP2 | frameshift deletion | HC LoF variant |

| | | | |
|---|---|---|---|
| chr13-100268784-G-A | PCCA | splicing | HC LoF variant |
| chr13-100515495-G-GACGGC( | PCCA | frameshift insertion | HC LoF variant |
| chr13-114324951-G-GATGATG | CHAMP1 | stopgain | HC LoF variant |
| chr14-21363130-T-TC | SUPT16H | frameshift insertion | HC LoF variant |
| chr14-21363133-T-TATATATTT( | SUPT16H | frameshift insertion | HC LoF variant |
| chr14-21409849-A-G | CHD8 | splicing | HC LoF variant |
| chr14-21429232-A-C | CHD8 | stopgain | HC LoF variant |
| chr14-31069841-A-G | AP4S1 | splicing | HC LoF variant |
| chr14-50201039-T-TCCTCTTA( | SOS2 | frameshift insertion | HC LoF variant |
| chr14-50201040-G-GT | SOS2 | frameshift insertion | HC LoF variant |
| chr14-50201043-C-CACAAGAT | SOS2 | stopgain | HC LoF variant |
| chr14-50201046-T-TCACGC | SOS2 | frameshift insertion | HC LoF variant |
| chr14-77027420-GC-G | IRF2BPL | frameshift deletion | HC LoF variant |
| chr14-77027422-TGCTGC-T | IRF2BPL | frameshift deletion | HC LoF variant |
| chr14-77027444-GC-G | IRF2BPL | frameshift deletion | HC LoF variant |
| chr14-77027446-TGCTGTTGC | IRF2BPL | frameshift deletion | HC LoF variant |
| chr14-77027446-TGCTGTTGC | IRF2BPL | frameshift deletion | HC LoF variant |
| chr14-77027450-G-GC | IRF2BPL | frameshift insertion | HC LoF variant |
| chr14-77027451-T-TGTTGC | IRF2BPL | frameshift insertion | HC LoF variant |
| chr14-77027451-T-TGC | IRF2BPL | frameshift insertion | HC LoF variant |
| chr14-77027487-CGCGGCGG( | IRF2BPL | frameshift deletion | HC LoF variant |
| chr14-77027489-CGGCGGCG( | IRF2BPL | frameshift deletion | HC LoF variant |
| chr14-77027495-CGG-C | IRF2BPL | frameshift deletion | HC LoF variant |
| chr14-77027498-CGGCG-C | IRF2BPL | frameshift deletion | HC LoF variant |
| chr14-78966168-G-GC | NRXN3 | frameshift insertion | HC LoF variant |
| chr14-78966169-A-AATCACCA | NRXN3 | stopgain | HC LoF variant |
| chr14-99510264-CCTGTGCAC | CCNK | frameshift deletion | HC LoF variant |
| chr14-99510270-CACCAGCCA | CCNK | frameshift deletion | HC LoF variant |
| chr14-102012104-TGCTGAGA- | DYNC1H1 | frameshift deletion | HC LoF variant |
| chr14-102030232-TC-T | DYNC1H1 | frameshift deletion | HC LoF variant |
| chr14-105382060-T-C | PACS2 | splicing | HC LoF variant |

| | | | |
|---|---|---|---|
| chr15-25375681-C-A | UBE3A | stopgain | HC LoF variant |
| chr15-28124153-C-A | HERC2 | stopgain | HC LoF variant |
| chr15-28201512-G-GT | HERC2 | frameshift insertion | HC LoF variant |
| chr15-28246761-A-ACCCCC | HERC2 | frameshift insertion | HC LoF variant |
| chr15-28246763-TGTAAG-T | HERC2 | frameshift deletion | HC LoF variant |
| chr15-28321414-CAG-C | HERC2 | frameshift deletion | HC LoF variant |
| chr15-43015841-C-CA | UBR1 | frameshift insertion | HC LoF variant |
| chr15-43015842-G-GCACCCG | UBR1 | frameshift insertion | HC LoF variant |
| chr15-43036593-C-CA | UBR1 | frameshift insertion | HC LoF variant |
| chr15-43056437-A-ACTGTTCC | UBR1 | stopgain | HC LoF variant |
| chr15-43527890-G-A | MAP1A | stopgain | HC LoF variant |
| chr15-43529173-A-AC | MAP1A | frameshift insertion | HC LoF variant |
| chr15-43529182-CAT-C | MAP1A | frameshift deletion | HC LoF variant |
| chr15-43529185-C-CAA | MAP1A | frameshift insertion | HC LoF variant |
| chr15-50930896-A-ACATC | AP4E1 | frameshift insertion | HC LoF variant |
| chr15-50949924-G-GCCCTCTA | AP4E1 | frameshift insertion | HC LoF variant |
| chr15-50949928-A-AGCCTCTC | AP4E1 | frameshift insertion | HC LoF variant |
| chr15-50958794-G-GGTAA | AP4E1 | frameshift insertion | HC LoF variant |
| chr15-50993574-GCTGGCATC | AP4E1 | frameshift deletion | HC LoF variant |
| chr15-56093427-C-CCCAGATC | RFX7 | frameshift insertion | HC LoF variant |
| chr15-63654163-GA-G | HERC1 | frameshift deletion | HC LoF variant |
| chr15-63678353-CACAAATTTT | HERC1 | frameshift deletion | HC LoF variant |
| chr15-63678366-C-G | HERC1 | splicing | HC LoF variant |
| chr15-63727743-AACTC-A | HERC1 | frameshift deletion | HC LoF variant |
| chr15-92991490-A-AAATT | CHD2 | stopgain | HC LoF variant |
| chr15-100580263-CT-C | LINS1 | frameshift deletion | HC LoF variant |
| chr15-100885359-A-ATGCAAT | ALDH1A3 | frameshift insertion | HC LoF variant |
| chr15-100885365-A-AAT | ALDH1A3 | frameshift insertion | HC LoF variant |
| chr15-100885366-G-GCA | ALDH1A3 | frameshift insertion | HC LoF variant |
| chr15-100905688-G-GT | ALDH1A3 | splicing | HC LoF variant |
| chr16-1505603-T-C | TELO2 | splicing | HC LoF variant |

| | | | |
|---|---|---|---|
| chr16-2082439-CCT-C | TSC2 | frameshift deletion | HC LoF variant |
| chr16-2083751-CCCCCAGGGT | TSC2 | frameshift deletion | HC LoF variant |
| chr16-2087923-TCCCTGCAGT | TSC2 | nonframeshift deletion | HC LoF variant |
| chr16-2087923-T-TCCCTGCAC | TSC2 | frameshift insertion | HC LoF variant |
| chr16-2770602-A-T | SRRM2 | splicing | HC LoF variant |
| chr16-3757373-C-A | CREBBP | stopgain | HC LoF variant |
| chr16-5073023-T-TGAAAAATG | ALG1 | frameshift insertion | HC LoF variant |
| chr16-5073152-G-T | ALG1 | splicing | HC LoF variant |
| chr16-5077443-A-T | ALG1 | splicing | HC LoF variant |
| chr16-8894579-A-ATTAGTAGA | USP7 | stopgain | HC LoF variant |
| chr16-30979988-G-GCCCCCC | SETD1A | frameshift insertion | HC LoF variant |
| chr16-30979991-G-GCCCCC | SETD1A | frameshift insertion | HC LoF variant |
| chr16-30979994-C-CA | SETD1A | frameshift insertion | HC LoF variant |
| chr16-30979994-C-CAA | SETD1A | frameshift insertion | HC LoF variant |
| chr16-72788687-G-GCT | ZFHX3 | frameshift insertion | HC LoF variant |
| chr16-72788692-G-GA | ZFHX3 | frameshift insertion | HC LoF variant |
| chr16-72957814-C-CCGCCGC | ZFHX3 | frameshift insertion | HC LoF variant |
| chr16-72957816-A-AGCCG | ZFHX3 | frameshift insertion | HC LoF variant |
| chr16-89282220-A-AC | ANKRD11 | frameshift insertion | HC LoF variant |
| chr16-89282225-CT-C | ANKRD11 | frameshift deletion | HC LoF variant |
| chr17-7417675-T-TG | NLGN2 | frameshift insertion | HC LoF variant |
| chr17-7417679-GCCACC-G | NLGN2 | frameshift deletion | HC LoF variant |
| chr17-7417679-G-GGCCCC | NLGN2 | frameshift insertion | HC LoF variant |
| chr17-7417679-G-GGCCC | NLGN2 | frameshift insertion | HC LoF variant |
| chr17-7417679-G-GGC | NLGN2 | frameshift insertion | HC LoF variant |
| chr17-7417681-CACCG-C | NLGN2 | frameshift deletion | HC LoF variant |
| chr17-7417681-CA-C | NLGN2 | frameshift deletion | HC LoF variant |
| chr17-7417684-CG-C | NLGN2 | frameshift deletion | HC LoF variant |
| chr17-7417684-CGCCA-C | NLGN2 | frameshift deletion | HC LoF variant |
| chr17-7417687-CA-C | NLGN2 | frameshift deletion | HC LoF variant |
| chr17-7417693-CACCG-C | NLGN2 | frameshift deletion | HC LoF variant |

| | | | |
|---|---|---|---|
| chr17-7417703-CT-C | NLGN2 | frameshift deletion | HC LoF variant |
| chr17-7417707-CTTCAT-C | NLGN2 | frameshift deletion | HC LoF variant |
| chr17-7417715-CTTCGGGCCC | NLGN2 | frameshift deletion | HC LoF variant |
| chr17-7417724-CTT-C | NLGN2 | frameshift deletion | HC LoF variant |
| chr17-7513774-CCA-C | POLR2A | splicing | HC LoF variant |
| chr17-7846892-CA-C | KDM6B | frameshift deletion | HC LoF variant |
| chr17-7846896-ACCCCT-A | KDM6B | frameshift deletion | HC LoF variant |
| chr17-7847694-CT-C | KDM6B | frameshift deletion | HC LoF variant |
| chr17-7847697-CA-C | KDM6B | frameshift deletion | HC LoF variant |
| chr17-7847704-C-CCCGG | KDM6B | frameshift insertion | HC LoF variant |
| chr17-7890881-GTGCGGCCTC | CHD3 | nonframeshift deletion | HC LoF variant |
| chr17-17793784-AGC-A | RAI1 | frameshift deletion | HC LoF variant |
| chr17-17793787-AG-A | RAI1 | frameshift deletion | HC LoF variant |
| chr17-17793787-AGCAG-A | RAI1 | frameshift deletion | HC LoF variant |
| chr17-17793787-AGCAGCAGC | RAI1 | frameshift deletion | HC LoF variant |
| chr17-17793787-AGCAGCAGC | RAI1 | frameshift deletion | HC LoF variant |
| chr17-17793787-AGCAGCAGC | RAI1 | frameshift deletion | HC LoF variant |
| chr17-30971327-AC-A | RNF135 | frameshift deletion | HC LoF variant |
| chr17-30971395-TC-T | RNF135 | frameshift deletion | HC LoF variant |
| chr17-30997307-AG-A | RNF135 | frameshift deletion | HC LoF variant |
| chr17-30998905-AG-A | RNF135 | frameshift deletion | HC LoF variant |
| chr17-31232072-G-T | NF1 | splicing | HC LoF variant |
| chr17-46094690-G-C | KANSL1 | stopgain | HC LoF variant |
| chr17-46171364-A-AC | KANSL1 | frameshift insertion | HC LoF variant |
| chr17-48075078-G-GTAACCGC | CBX1 | frameshift insertion | HC LoF variant |
| chr17-57979242-G-GC | VEZF1 | frameshift insertion | HC LoF variant |
| chr17-57979243-T-TGCTGCTG | VEZF1 | frameshift insertion | HC LoF variant |
| chr17-57979245-GC-G | VEZF1 | frameshift deletion | HC LoF variant |
| chr17-57979247-TGC-T | VEZF1 | frameshift deletion | HC LoF variant |
| chr17-57979247-TGCTGC-T | VEZF1 | frameshift deletion | HC LoF variant |
| chr17-60647902-G-A | PPM1D | stopgain | HC LoF variant |

| | | | |
|---|---|---|---|
| chr17-61965401-T-TCA | MED13 | frameshift insertion | HC LoF variant |
| chr17-61965402-G-GGTGTTG( | MED13 | frameshift insertion | HC LoF variant |
| chr17-62612534-CTG-C | TLK2 | frameshift deletion | HC LoF variant |
| chr17-62612538-G-GAA | TLK2 | frameshift insertion | HC LoF variant |
| chr17-67912039-T-TCATCA | BPTF | frameshift insertion | HC LoF variant |
| chr17-67959666-A-AGC | BPTF | frameshift insertion | HC LoF variant |
| chr17-80213850-TCGGGCTGC | SGSH | frameshift deletion | HC LoF variant |
| chr18-33739485-T-TGGTTG | ASXL3 | frameshift insertion | HC LoF variant |
| chr18-33745913-C-CG | ASXL3 | frameshift insertion | HC LoF variant |
| chr18-33745919-C-CG | ASXL3 | frameshift insertion | HC LoF variant |
| chr18-33745932-C-CCGGGGG | ASXL3 | frameshift insertion | HC LoF variant |
| chr18-36142348-G-A | ELP2 | splicing | HC LoF variant |
| chr18-36164649-A-AGCTG | ELP2 | frameshift insertion | HC LoF variant |
| chr18-45063472-T-TG | SETBP1 | frameshift insertion | HC LoF variant |
| chr18-45063478-CG-C | SETBP1 | frameshift deletion | HC LoF variant |
| chr18-45063478-CGCCACCGC | SETBP1 | frameshift deletion | HC LoF variant |
| chr18-45063495-CTG-C | SETBP1 | frameshift deletion | HC LoF variant |
| chr18-45063497-G-GGC | SETBP1 | frameshift insertion | HC LoF variant |
| chr18-45063502-CACCGCCG- | SETBP1 | frameshift deletion | HC LoF variant |
| chr18-45063519-CTG-C | SETBP1 | frameshift deletion | HC LoF variant |
| chr18-47046493-CT-C | KATNAL2 | frameshift deletion | HC LoF variant |
| chr18-62072726-C-T | PIGN | splicing | HC LoF variant |
| chr18-62088797-G-A | PIGN | stopgain | HC LoF variant |
| chr18-75285870-G-GCGCCCC | TSHZ1 | frameshift insertion | HC LoF variant |
| chr18-75285870-G-GGC | TSHZ1 | frameshift insertion | HC LoF variant |
| chr18-75285870-G-GGCCC | TSHZ1 | frameshift insertion | HC LoF variant |
| chr18-75285870-G-GGCCCCC | TSHZ1 | frameshift insertion | HC LoF variant |
| chr18-75285871-C-CG | TSHZ1 | frameshift insertion | HC LoF variant |
| chr18-75285875-CA-C | TSHZ1 | frameshift deletion | HC LoF variant |
| chr18-75285893-CTG-C | TSHZ1 | frameshift deletion | HC LoF variant |
| chr18-75285895-G-GGCCCCC | TSHZ1 | frameshift insertion | HC LoF variant |

| | | | |
|---|---|---|---|
| chr18-75285899-CGTCAGCA-( | TSHZ1 | frameshift deletion | HC LoF variant |
| chr18-75285899-CGT-C | TSHZ1 | frameshift deletion | HC LoF variant |
| chr18-75285902-CAG-C | TSHZ1 | frameshift deletion | HC LoF variant |
| chr18-75285908-CACTGG-C | TSHZ1 | frameshift deletion | HC LoF variant |
| chr19-11033765-A-C | SMARCA4 | splicing | HC LoF variant |
| chr19-11033766-GAGCAGAC-( | SMARCA4 | frameshift deletion | HC LoF variant |
| chr19-13135433-C-T | NACC1 | stopgain | HC LoF variant |
| chr19-13207336-C-A | CACNA1A | stopgain | HC LoF variant |
| chr19-13207438-AAGGCG-A | CACNA1A | frameshift deletion | HC LoF variant |
| chr19-13913299-TAAAG-T | CC2D1A | nonframeshift deletion | HC LoF variant |
| chr19-13918778-T-TCGCAGC( | CC2D1A | frameshift insertion | HC LoF variant |
| chr19-13923385-G-GT | CC2D1A | frameshift insertion | HC LoF variant |
| chr19-13923387-C-CAG | CC2D1A | frameshift insertion | HC LoF variant |
| chr19-13926820-G-A | CC2D1A | splicing | HC LoF variant |
| chr19-15256126-G-GTGAACT( | BRD4 | frameshift insertion | HC LoF variant |
| chr19-17606183-CATGTGGA-( | UNC13A | frameshift deletion | HC LoF variant |
| chr19-35720465-A-ATTGAGGA | KMT2B | stopgain | HC LoF variant |
| chr19-35720589-T-TG | KMT2B | frameshift insertion | HC LoF variant |
| chr19-35720606-CTT-C | KMT2B | frameshift deletion | HC LoF variant |
| chr19-35721215-CAG-C | KMT2B | frameshift deletion | HC LoF variant |
| chr19-35721221-CT-C | KMT2B | frameshift deletion | HC LoF variant |
| chr19-35721233-CGG-C | KMT2B | frameshift deletion | HC LoF variant |
| chr19-41975740-C-CG | ATP1A3 | frameshift insertion | HC LoF variant |
| chr19-41975743-TC-T | ATP1A3 | frameshift deletion | HC LoF variant |
| chr19-42295141-C-CCG | CIC | frameshift insertion | HC LoF variant |
| chr19-42295142-A-AGG | CIC | frameshift insertion | HC LoF variant |
| chr19-42295142-A-AG | CIC | frameshift insertion | HC LoF variant |
| chr19-42295143-G-GGC | CIC | frameshift insertion | HC LoF variant |
| chr19-42295143-G-GGCCC | CIC | frameshift insertion | HC LoF variant |
| chr19-42295143-G-GGGCCCC | CIC | frameshift insertion | HC LoF variant |
| chr19-45771770-C-T | DM1-AS;DMPK | splicing | HC LoF variant |

| | | | |
|---|---|---|---|
| chr19-47483381-T-C | KPTN | splicing | HC LoF variant |
| chr19-47695362-CAGG-C | BICRA | frameshift deletion | HC LoF variant |
| chr19-47695363-A-C | BICRA | splicing | HC LoF variant |
| chr19-47695364-G-C | BICRA | splicing | HC LoF variant |
| chr19-47699405-G-GGTGAGA | BICRA | frameshift insertion | HC LoF variant |
| chr19-49596842-C-CA | PRR12 | frameshift insertion | HC LoF variant |
| chr19-50668945-GT-G | SHANK1 | frameshift deletion | HC LoF variant |
| chr19-50713950-C-CT | SHANK1 | splicing | HC LoF variant |
| chr2-24726700-C-A | NCOA1 | stopgain | HC LoF variant |
| chr2-24768341-A-AC | NCOA1 | frameshift insertion | HC LoF variant |
| chr2-25239134-T-TCATACCGG | DNMT3A | stopgain | HC LoF variant |
| chr2-25240326-CTT-C | DNMT3A | frameshift deletion | HC LoF variant |
| chr2-25244153-A-C | DNMT3A | splicing | HC LoF variant |
| chr2-25246158-A-G | DNMT3A | splicing | HC LoF variant |
| chr2-25246159-C-A | DNMT3A | splicing | HC LoF variant |
| chr2-47832434-A-AATGTAAAA | FBXO11 | stopgain | HC LoF variant |
| chr2-54659952-G-GGAAGGGA | SPTBN1 | stopgain | HC LoF variant |
| chr2-144398571-T-TTGATTATC | ZEB2 | frameshift insertion | HC LoF variant |
| chr2-156326884-C-CCTTCGAT | NR4A2 | frameshift insertion | HC LoF variant |
| chr2-165297059-A-AATACTATA | SCN2A | frameshift insertion | HC LoF variant |
| chr2-165314098-A-ATCAAATT | SCN2A | stopgain | HC LoF variant |
| chr2-165331511-T-TGCTAGAA | SCN2A | stopgain | HC LoF variant |
| chr2-166058593-T-TAAATATC | SCN1A | frameshift insertion | HC LoF variant |
| chr2-175992350-TA-T | LNPK | stopgain | HC LoF variant |
| chr2-175992353-TCTTCCAAC | LNPK | frameshift deletion | HC LoF variant |
| chr2-178527146-C-A | TTN | stopgain | HC LoF variant |
| chr2-178528773-G-A | TTN | stopgain | HC LoF variant |
| chr2-178528775-T-TTCATTCAT | TTN | stopgain | HC LoF variant |
| chr2-178530083-C-CCA | TTN | frameshift insertion | HC LoF variant |
| chr2-178531632-C-A | TTN | stopgain | HC LoF variant |
| chr2-178539132-C-A | TTN | stopgain | HC LoF variant |

| | | | |
|---|---|---|---|
| chr2-178540208-GT-G | TTN | frameshift deletion | HC LoF variant |
| chr2-178544199-A-G | TTN | splicing | HC LoF variant |
| chr2-178549342-T-TGCTCAGC | TTN | frameshift insertion | HC LoF variant |
| chr2-178549347-T-TGAATA | TTN | frameshift insertion | HC LoF variant |
| chr2-178549348-C-CTAGTAAT | TTN | frameshift insertion | HC LoF variant |
| chr2-178551799-ATTTC-A | TTN | frameshift deletion | HC LoF variant |
| chr2-178559368-C-A | TTN | stopgain | HC LoF variant |
| chr2-178564082-T-TATGCTCC | TTN | frameshift insertion | HC LoF variant |
| chr2-178568947-T-A | TTN | stopgain | HC LoF variant |
| chr2-178573140-C-CAGTACCT | TTN | stopgain | HC LoF variant |
| chr2-178573722-G-T | TTN | stopgain | HC LoF variant |
| chr2-178590118-ATT-A | TTN | frameshift deletion | HC LoF variant |
| chr2-178590670-C-CCTAAA | TTN | stopgain | HC LoF variant |
| chr2-178601788-C-T | TTN | splicing | HC LoF variant |
| chr2-178604085-T-TA | TTN | frameshift insertion | HC LoF variant |
| chr2-178624464-C-A | TTN | splicing | HC LoF variant |
| chr2-178645992-T-TTTAGGCC | TTN | frameshift insertion | HC LoF variant |
| chr2-178654266-TA-T | TTN | frameshift deletion | HC LoF variant |
| chr2-178665777-G-A | TTN | stopgain | HC LoF variant |
| chr2-178683210-C-A | TTN | splicing | HC LoF variant |
| chr2-178701170-AGG-A | TTN | frameshift deletion | HC LoF variant |
| chr2-178704626-ATT-A | TTN | frameshift deletion | HC LoF variant |
| chr2-178712594-C-A | TTN | splicing | HC LoF variant |
| chr2-178719553-C-G | TTN | splicing | HC LoF variant |
| chr2-178720256-G-A | TTN | stopgain | HC LoF variant |
| chr2-178723855-C-T | TTN | splicing | HC LoF variant |
| chr2-178731972-C-A | TTN | splicing | HC LoF variant |
| chr2-178789977-C-A | TTN | splicing | HC LoF variant |
| chr2-178790707-C-T | TTN | splicing | HC LoF variant |
| chr2-178792199-T-C | TTN | splicing | HC LoF variant |
| chr2-178793425-C-CTAGACAA | TTN | frameshift insertion | HC LoF variant |

| | | | |
|---|---|---|---|
| chr2-182925777-TTCCAA-T | NCKAP1 | frameshift deletion | HC LoF variant |
| chr2-182962184-G-GCTTAGTC | NCKAP1 | frameshift insertion | HC LoF variant |
| chr2-196318961-G-GGAGGAA | HECW2 | frameshift insertion | HC LoF variant |
| chr2-218812299-CG-C | CYP27A1 | frameshift deletion | HC LoF variant |
| chr2-218814188-G-A | CYP27A1 | splicing | HC LoF variant |
| chr2-227366149-CAT-C | TM4SF20 | frameshift deletion | HC LoF variant |
| chr2-229778432-C-CA | TRIP12 | splicing | HC LoF variant |
| chr2-229778434-A-AT | TRIP12 | frameshift insertion | HC LoF variant |
| chr2-229813920-GAA-G | TRIP12 | frameshift deletion | HC LoF variant |
| chr2-229858950-G-GAGGAGA | TRIP12 | frameshift insertion | HC LoF variant |
| chr2-229859293-G-T | TRIP12 | stopgain | HC LoF variant |
| chr2-232794829-T-TCC | GIGYF2 | frameshift insertion | HC LoF variant |
| chr2-232794831-C-CTAAGCAC | GIGYF2 | stopgain | HC LoF variant |
| chr2-232794833-C-CTT | GIGYF2 | frameshift insertion | HC LoF variant |
| chr2-232847515-C-CCG | GIGYF2 | frameshift insertion | HC LoF variant |
| chr2-232847517-A-AG | GIGYF2 | frameshift insertion | HC LoF variant |
| chr20-32158450-G-T | TM9SF4 | splicing | HC LoF variant |
| chr20-38574151-TC-T | RALGAPB | frameshift deletion | HC LoF variant |
| chr20-38574154-ACAACTCTT- | RALGAPB | frameshift deletion | HC LoF variant |
| chr20-62064972-AGAGTGGCG | TAF4 | frameshift deletion | HC LoF variant |
| chr20-62064992-TGGCGGGG( | TAF4 | frameshift deletion | HC LoF variant |
| chr21-33554085-T-TTC | SON | frameshift insertion | HC LoF variant |
| chr21-33576378-G-GA | SON | frameshift insertion | HC LoF variant |
| chr21-33576390-GA-G | SON | frameshift deletion | HC LoF variant |
| chr21-37493137-G-T | DYRK1A | Stop gain | HC LoF variant |
| chr21-37511973-TGTTGA-T | DYRK1A | frameshift deletion | HC LoF variant |
| chr21-37511981-CTCATCCTGT | DYRK1A | frameshift deletion | HC LoF variant |
| chr21-46558242-CAT-C | DIP2A | frameshift deletion | HC LoF variant |
| chr21-46567430-G-GTGCACTA | DIP2A | frameshift insertion | HC LoF variant |
| chr21-46567432-A-AATATTGAT | DIP2A | stopgain | HC LoF variant |
| chr22-20983039-C-CA | LZTR1 | frameshift insertion | HC LoF variant |

| | | | |
|---|---|---|---|
| chr22-20987503-G-C | LZTR1 | splicing | HC LoF variant |
| chr22-20990526-G-GT | LZTR1 | splicing | HC LoF variant |
| chr22-20994728-G-C | LZTR1 | splicing | HC LoF variant |
| chr22-20995835-C-CT | LZTR1 | frameshift insertion | HC LoF variant |
| chr22-20995837-G-GGAGGTG | LZTR1 | frameshift insertion | HC LoF variant |
| chr22-31792102-GGTGA-G | DEPDC5 | splicing | HC LoF variant |
| chr22-40349868-G-T | ADSL | stopgain | HC LoF variant |
| chr22-49913518-C-T | ALG12 | splicing | HC LoF variant |
| chr22-50582324-C-CAGGGTC/ | CHKB | frameshift insertion | HC LoF variant |
| chr22-50674573-CCG-C | SHANK3 | splicing | HC LoF variant |
| chr22-50697540-CTGTATTCG/ | SHANK3 | splicing | HC LoF variant |
| chr22-50697555-A-C | SHANK3 | splicing | HC LoF variant |
| chr22-50697556-G-GGCCCCG | SHANK3 | frameshift deletion | HC LoF variant |
| chr22-50697556-G-GCGGGCC | SHANK3 | frameshift insertion | HC LoF variant |
| chr3-1295660-C-T | CNTN6 | stopgain | HC LoF variant |
| chr3-1352452-G-T | CNTN6 | splicing | HC LoF variant |
| chr3-1383176-G-GGTAA | CNTN6 | frameshift insertion | HC LoF variant |
| chr3-1403346-AG-A | CNTN6 | frameshift deletion | HC LoF variant |
| chr3-4676695-T-TAA | ITPR1 | frameshift insertion | HC LoF variant |
| chr3-9447306-G-GA | SETD5 | frameshift insertion | HC LoF variant |
| chr3-9447307-G-GTCTCTGAG | SETD5 | frameshift insertion | HC LoF variant |
| chr3-14148846-C-A | XPC | stopgain | HC LoF variant |
| chr3-14158640-G-A | XPC | stopgain | HC LoF variant |
| chr3-14164836-G-A | XPC | stopgain | HC LoF variant |
| chr3-14170468-CT-C | XPC | frameshift deletion | HC LoF variant |
| chr3-47122611-T-TATGA | SETD2 | frameshift insertion | HC LoF variant |
| chr3-47122612-C-CTGGCACC | SETD2 | frameshift insertion | HC LoF variant |
| chr3-54581865-GACAA-G | CACNA2D3 | frameshift deletion | HC LoF variant |
| chr3-54918482-GCT-G | LRTM1 | frameshift deletion | HC LoF variant |
| chr3-54918600-CA-C | LRTM1 | frameshift deletion | HC LoF variant |
| chr3-54925031-AT-A | LRTM1 | frameshift deletion | HC LoF variant |

| | | | |
|---|---|---|---|
| chr3-64022399-A-ACTTCTCAC | PSMD6 | stopgain | HC LoF variant |
| chr3-136421163-C-CGTTGTTA | STAG1 | stopgain | HC LoF variant |
| chr3-136521393-T-TACAA | STAG1 | frameshift insertion | HC LoF variant |
| chr3-151188481-G-A | MED12L | splicing | HC LoF variant |
| chr3-151294305-T-TGCTCACC | GPR87 | stopgain | HC LoF variant |
| chr3-151294668-G-T | GPR87 | stopgain | HC LoF variant |
| chr3-151338475-TG-T | P2RY12 | frameshift deletion | HC LoF variant |
| chr3-151372635-C-T | MED12L | stopgain | HC LoF variant |
| chr3-158203246-G-GATTGTAT | RSRC1 | splicing | HC LoF variant |
| chr3-184244686-A-AAG | ALG3 | frameshift insertion | HC LoF variant |
| chr4-48565039-A-AATGTAGGA | FRYL | frameshift insertion | HC LoF variant |
| chr4-48570881-A-ATGTATGTG | FRYL | stopgain | HC LoF variant |
| chr4-48594017-CT-C | FRYL | splicing | HC LoF variant |
| chr4-82356630-C-CAA | HNRNPD | frameshift insertion | HC LoF variant |
| chr4-84705512-C-A | WDFY3 | splicing | HC LoF variant |
| chr4-106244657-G-A | TBCK | stopgain | HC LoF variant |
| chr4-106308767-C-A | TBCK | splicing | HC LoF variant |
| chr4-113354556-A-T | ANK2 | stopgain | HC LoF variant |
| chr4-113359220-C-CTGTGTCA | ANK2 | frameshift insertion | HC LoF variant |
| chr4-113369636-CCCCAACA-C | ANK2 | frameshift deletion | HC LoF variant |
| chr4-113369647-AGCGAGCGC | ANK2 | frameshift deletion | HC LoF variant |
| chr4-113369662-TCTCCC-T | ANK2 | frameshift deletion | HC LoF variant |
| chr4-113369669-TCA-T | ANK2 | frameshift deletion | HC LoF variant |
| chr4-113369677-GAACCCGAA | ANK2 | frameshift deletion | HC LoF variant |
| chr4-113369689-C-CTGGTTTT | ANK2 | frameshift insertion | HC LoF variant |
| chr4-113369691-CTCAGA-C | ANK2 | frameshift deletion | HC LoF variant |
| chr4-113369698-CACAGAGA-C | ANK2 | frameshift deletion | HC LoF variant |
| chr4-145110967-G-T | ABCE1 | splicing | HC LoF variant |
| chr4-148120287-CTCTG-C | NR3C2 | frameshift deletion | HC LoF variant |
| chr5-14487813-CG-C | TRIO | frameshift deletion | HC LoF variant |
| chr5-61332763-CGG-C | ZSWIM6 | frameshift deletion | HC LoF variant |

| | | | |
|---|---|---|---|
| chr5-61332766-CCGCAACCTC | ZSWIM6 | frameshift deletion | HC LoF variant |
| chr5-61332769-CAACCT-C | ZSWIM6 | frameshift deletion | HC LoF variant |
| chr5-61332772-CCTCG-C | ZSWIM6 | frameshift deletion | HC LoF variant |
| chr5-61332776-G-GCC | ZSWIM6 | frameshift insertion | HC LoF variant |
| chr5-61538938-G-GCCATTTCC | ZSWIM6 | frameshift insertion | HC LoF variant |
| chr5-61544281-G-GAACA | ZSWIM6 | frameshift insertion | HC LoF variant |
| chr6-26104135-T-TA | H4C3 | frameshift insertion | HC LoF variant |
| chr6-26104247-C-CT | H4C3 | frameshift insertion | HC LoF variant |
| chr6-26156602-A-AT | H1-4 | stopgain | HC LoF variant |
| chr6-33451829-G-GGCCCC | SYNGAP1 | frameshift insertion | HC LoF variant |
| chr6-33451836-CAG-C | SYNGAP1 | frameshift deletion | HC LoF variant |
| chr6-33451838-G-GCCCCCCC | SYNGAP1 | frameshift insertion | HC LoF variant |
| chr6-63685659-A-ATGAG | PHF3 | frameshift insertion | HC LoF variant |
| chr6-63712801-CCTTACAGGA | PHF3 | frameshift deletion | HC LoF variant |
| chr6-63720676-G-GA | EYS | frameshift insertion | HC LoF variant |
| chr6-63721281-A-T | EYS | stopgain | HC LoF variant |
| chr6-72400534-AT-A | RIMS1 | frameshift deletion | HC LoF variant |
| chr6-72400538-T-TCTCCAATT | RIMS1 | stopgain | HC LoF variant |
| chr6-87260588-A-AAATGAGC | ZNF292 | frameshift insertion | HC LoF variant |
| chr6-135404950-CT-C | AHI1 | frameshift deletion | HC LoF variant |
| chr6-135427196-GGCTCATTT- | AHI1 | frameshift deletion | HC LoF variant |
| chr6-135457594-G-A | AHI1 | stopgain | HC LoF variant |
| chr6-156778291-A-AGC | ARID1B | frameshift insertion | HC LoF variant |
| chr6-156778292-A-AGCAG | ARID1B | frameshift insertion | HC LoF variant |
| chr6-156778292-A-AG | ARID1B | frameshift insertion | HC LoF variant |
| chr6-156778292-A-AGCAGCAC | ARID1B | frameshift insertion | HC LoF variant |
| chr7-1473588-G-A | INTS1 | stopgain | HC LoF variant |
| chr7-1476659-T-C | INTS1 | splicing | HC LoF variant |
| chr7-1497161-G-A | INTS1 | stopgain | HC LoF variant |
| chr7-44229442-C-A | CAMK2B | stopgain | HC LoF variant |
| chr7-98910216-C-CA | TRRAP | frameshift insertion | HC LoF variant |

| | | | |
|---|---|---|---|
| chr7-98910216-C-CG | TRRAP | frameshift insertion | HC LoF variant |
| chr7-98910217-T-TG | TRRAP | frameshift insertion | HC LoF variant |
| chr7-98910218-G-GGCCC | TRRAP | frameshift insertion | HC LoF variant |
| chr7-98910218-G-GGC | TRRAP | frameshift insertion | HC LoF variant |
| chr7-98910218-G-GCCCCCCC | TRRAP | frameshift insertion | HC LoF variant |
| chr7-98910222-CACCTCCA-C | TRRAP | frameshift deletion | HC LoF variant |
| chr7-98910222-CACCT-C | TRRAP | frameshift deletion | HC LoF variant |
| chr7-98910222-CA-C | TRRAP | frameshift deletion | HC LoF variant |
| chr7-98910225-CT-C | TRRAP | frameshift deletion | HC LoF variant |
| chr7-98910246-CACCTG-C | TRRAP | frameshift deletion | HC LoF variant |
| chr7-98910249-CTG-C | TRRAP | frameshift deletion | HC LoF variant |
| chr7-98910258-CTGTGA-C | TRRAP | frameshift deletion | HC LoF variant |
| chr7-98910267-CGG-C | TRRAP | frameshift deletion | HC LoF variant |
| chr7-100101262-A-G | MCM7 | splicing | HC LoF variant |
| chr7-100655848-GCC-G | ACTL6B | frameshift deletion | HC LoF variant |
| chr7-103523529-G-GCAAA | RELN | frameshift insertion | HC LoF variant |
| chr7-103545183-G-T | RELN | stopgain | HC LoF variant |
| chr7-105112789-C-CCG | KMT2E | frameshift insertion | HC LoF variant |
| chr7-105112792-CA-C | KMT2E | frameshift deletion | HC LoF variant |
| chr7-105112807-CTGGT-C | KMT2E | frameshift deletion | HC LoF variant |
| chr7-114629931-C-T | FOXP2 | stopgain | HC LoF variant |
| chr7-114631524-CCAG-C | FOXP2 | splicing | HC LoF variant |
| chr7-117718045-ATTAATAGTT( | CTTNBP2 | stopgain | HC LoF variant |
| chr7-117718094-CTTAAATCAT | CTTNBP2 | frameshift deletion | HC LoF variant |
| chr7-117735342-CAG-C | CTTNBP2 | frameshift deletion | HC LoF variant |
| chr7-147562135-TAG-T | CNTNAP2 | splicing | HC LoF variant |
| chr7-147562140-ATC-A | CNTNAP2 | frameshift deletion | HC LoF variant |
| chr7-147562144-ACG-A | CNTNAP2 | stop gain | HC LoF variant |
| chr7-152162427-T-TATGACTG | KMT2C | stopgain | HC LoF variant |
| chr7-152177898-A-AGCTTGCC | KMT2C | frameshift insertion | HC LoF variant |
| chr7-152178011-C-CT | KMT2C | splicing | HC LoF variant |

| | | | |
|---|---|---|---|
| chr7-152178011-C-A | KMT2C | splicing | HC LoF variant |
| chr7-152187302-G-T | KMT2C | stopgain | HC LoF variant |
| chr7-152194508-G-T | KMT2C | stopgain | HC LoF variant |
| chr7-152247986-G-GT | KMT2C | stopgain | HC LoF variant |
| chr8-6444589-G-GA | MCPH1 | frameshift insertion | HC LoF variant |
| chr8-6444766-AC-A | MCPH1 | frameshift deletion | HC LoF variant |
| chr8-6445283-G-T | MCPH1 | stopgain | HC LoF variant |
| chr8-15659579-G-GGAGGAAA | TUSC3 | frameshift insertion | HC LoF variant |
| chr8-15662155-G-C | TUSC3 | splicing | HC LoF variant |
| chr8-15806431-GA-G | TUSC3,MSR1 | frameshift deletion | HC LoF variant |
| chr8-33507246-C-A | TTI2 | stopgain | HC LoF variant |
| chr8-99778780-C-T | VPS13B | stopgain | HC LoF variant |
| chr8-99819925-G-T | VPS13B | stopgain | HC LoF variant |
| chr8-99832366-TAG-T | VPS13B | splicing | HC LoF variant |
| chr8-99832368-G-T | VPS13B | splicing | HC LoF variant |
| chr8-99832368-G-GTTTTTTTT | VPS13B | splicing | HC LoF variant |
| chr8-99832368-G-GTTTTTTTT | VPS13B | splicing | HC LoF variant |
| chr8-99832374-TCG-T | VPS13B | frameshift deletion | HC LoF variant |
| chr8-100706805-GCTAAAAAAT | PABPC1 | splicing | HC LoF variant |
| chr8-100706997-G-GGATGAG | PABPC1 | frameshift insertion | HC LoF variant |
| chr8-100706997-G-GGATGAG | PABPC1 | frameshift insertion | HC LoF variant |
| chr8-100709671-C-A | PABPC1 | stopgain | HC LoF variant |
| chr8-100709704-CT-C | PABPC1 | frameshift deletion | HC LoF variant |
| chr8-100712790-C-CT | PABPC1 | splicing | HC LoF variant |
| chr8-104013524-C-CTTCCTGA | RIMS2 | frameshift insertion | HC LoF variant |
| chr8-139885970-G-A | TRAPPC9 | splicing | HC LoF variant |
| chr9-2039817-A-AGC | SMARCA2 | frameshift insertion | HC LoF variant |
| chr9-2039818-A-AGCAG | SMARCA2 | frameshift insertion | HC LoF variant |
| chr9-2039818-A-AG | SMARCA2 | frameshift insertion | HC LoF variant |
| chr9-70536027-G-A | TRPM3 | stopgain | HC LoF variant |
| chr9-70784129-C-CTATTAATGA | TRPM3 | stopgain | HC LoF variant |

| | | | |
|---|---|---|---|
| chr9-106926840-CCTGA-C | ZNF462 | frameshift deletion | HC LoF variant |
| chr9-106926847-G-T | ZNF462 | stopgain | HC LoF variant |
| chr9-106927204-T-TC | ZNF462 | frameshift insertion | HC LoF variant |
| chr9-106927204-T-TGC | ZNF462 | frameshift insertion | HC LoF variant |
| chr9-106927204-T-TGCCCCC( | ZNF462 | frameshift insertion | HC LoF variant |
| chr9-106927217-CACAA-C | ZNF462 | frameshift deletion | HC LoF variant |
| chr9-106927219-CAA-C | ZNF462 | frameshift deletion | HC LoF variant |
| chr9-106927221-A-ACCCCCC( | ZNF462 | frameshift insertion | HC LoF variant |
| chr9-107011004-G-T | ZNF462 | stopgain | HC LoF variant |
| chr9-137744014-A-AGCAGGC( | EHMT1 | frameshift insertion | HC LoF variant |
| chrX-12720542-G-T | FRMPD4 | stopgain | HC LoF variant |
| chrX-17728277-A-AGT | NHS | frameshift insertion | HC LoF variant |
| chrX-17728279-CAG-C | NHS | frameshift deletion | HC LoF variant |
| chrX-21561555-G-GAAAC | CNKSR2 | frameshift insertion | HC LoF variant |
| chrX-21845198-A-AGGAAGAT( | MBTPS2 | stopgain | HC LoF variant |
| chrX-31323630-G-GGAATGTC. | DMD | frameshift insertion | HC LoF variant |
| chrX-32342182-G-GTTACT | DMD | stopgain | HC LoF variant |
| chrX-41216447-C-CTGCT | USP9X | frameshift insertion | HC LoF variant |
| chrX-48903064-C-T | PQBP1 | stopgain | HC LoF variant |
| chrX-63655515-C-CGGAACAC | ARHGEF9 | stopgain | HC LoF variant |
| chrX-71140822-C-CTT | MED12 | frameshift insertion | HC LoF variant |
| chrX-74740391-G-GATATATGA | NEXMIF | stopgain | HC LoF variant |
| chrX-77696669-T-TGGTTGTG | ATRX | frameshift insertion | HC LoF variant |
| chrX-80676956-C-CCTATTT | BRWD3 | stopgain | HC LoF variant |
| chrX-80745696-C-CTGTGT | BRWD3 | frameshift insertion | HC LoF variant |
| chrX-111142101-A-ATTGCTAAT | PAK3 | stopgain | HC LoF variant |
| chrX-134415106-C-CTTTCTAA | PHF6 | frameshift insertion | HC LoF variant |
| chrX-141879368-GCTACACCC | MAGEC3 | frameshift deletion | HC LoF variant |
| chrX-141881464-C-T | MAGEC3 | stopgain | HC LoF variant |
| chrX-147928713-A-ATT | FMR1 | frameshift insertion | HC LoF variant |
| chrX-147928716-G-T | FMR1 | stopgain | HC LoF variant |

| | | | |
|---|---|---|---|
| chrX-148958417-C-A | AFF2 | stopgain | HC LoF variant |

**Supplementary Table 3:** Highlighting the PVS1 assignment criteria, the satisfying variants (P/LP) are listed with their parameters.

| variant | gene | variant class | ACMG_Attribute | Sfari/Clingen | Clinvar >5% | MANE_Transcript | Auto PVS1 | Literature evidence |
|---|---|---|---|---|---|---|---|---|
| chr1-202753014-T-TTTGG | KDM5B | frameshift variant | PVS1 | DE | 92.96 | NM_006618.5 | Very Strong | PMID: 30409806:A Kdm5b loss-of-function mouse model shows subviability in homozygous null mice, with fully penetrant vertebral defects and neurobehavioral abnormalities. These findings support a loss-of-function mechanism consistent with PVS1. |
| chr13-35156147-T-TCCTTA | NBEA | frameshift variant | PVS1 | Definitive | 77.78 | NM_001385012.1 | Very Strong | PMID: 23153818:Haploinsufficiency of NBEA is supported by de novo LoF variants in ASD patients and Nbea+/− mouse models showing ASD-like behaviors and impaired neuroplasticity, justifying application of PVS1. |
| chr13-35593385-C-CTTCCTCCATCCCTTTATTTTTTAAGTAG | NBEA | frameshift variant | PVS1 | Definitive | 77.78 | NM_001385012.1 | Very Strong | PMID: 23153818:Haploinsufficiency of NBEA is supported by de novo LoF variants in ASD patients and Nbea+/− mouse models showing ASD-like behaviors and impaired neuroplasticity, justifying application of PVS1. |
| chr14-21429232-A-C | CHD8 | stop gained | PVS1 | Definitive | 86.19 | NM_001170629.2 | Very Strong | PMID: 27602517: CHD8 haploinsufficiency leads to ASD-like phenotypes in mouse models, supporting loss-of-function as a disease mechanism and application of PVS1. |
| chr15-92991490-A-AAATT | CHD2 | stop gained & frameshift variant | PVS1 | Definitive | 78.01 | NM_001271.4 | Very Strong | PMID: 30344048,PMID: 24834135:CHD2 haploinsufficiency causes neurodevelopmental and behavioral deficits in both humans and Chd2+/- mice, supporting loss-of-function as a disease mechanism and application of PVS1. |
| chr16-3757373-C-A | CREBBP | stop gained | PVS1 | Definitive | 71.64 | NM_004380.3 | Very Strong | PMID: 26730956:CBP CH1 domain loss in mice causes ASD-like traits, supporting CREBBP loss-of-function as a disease mechanism and application of PVS1. |
| chr16-89282220-A-AC | ANKRD11 | frameshift variant | PVS1 | Definitive | 93.11 | NM_013275.6 | Very Strong | PMID:29274743:ANKRD11 loss-of-function impairs neuronal development and epigenetic regulation, supporting its role in KBG syndrome and ASD, consistent with PVS1 application. |
| chr16-89282225-CT-C | ANKRD11 | frameshift variant | PVS1 | Definitive | 93.11 | NM_013275.6 | Very Strong | PMID:29274743:ANKRD11 loss-of-function impairs neuronal development and epigenetic regulation, supporting its role in KBG syndrome and ASD, consistent with PVS1 application. |
| chr17-31232072-G-T | NF1 | splice acceptor variant | PVS1 | Definitive | 85.83 | NM_001042492.3 | Very Strong | PMID: 32640222:Nf1 loss in Drosophila leads to social behavioral deficits, supporting NFI haploinsufficiency as a disease mechanism and application of PVS1. |
| chr21-37493137-G-T | DYRK1A | stop gained | PVS1 | Definitive | 82.85 | NM_001347721.2 | Very Strong | PMID: 30831192:Haploinsufficient Dyrk1a+/− mouse models replicate key human phenotypes, supporting loss-of-function as a disease mechanism and application of PVS1. |
| chr21-37511973-TGTTGA-T | DYRK1A | frameshift variant | PVS1 | Definitive | 82.85 | NM_001347721.2 | Strong | PMID: 30831192:Haploinsufficient Dyrk1a+/− mouse models replicate key human phenotypes, supporting loss-of-function as a disease mechanism and application of PVS1. |
| chr21-37511981-CTCATCCTGTTCAAG-C | DYRK1A | frameshift variant | PVS1 | Definitive | 82.85 | NM_001347721.2 | Strong | PMID: 30831192:Haploinsufficient Dyrk1a+/− mouse models replicate key human phenotypes, supporting loss-of-function as a disease mechanism and application of PVS1. |
| chr4-84705512-C-A | WDFY3 | splice acceptor variant | PVS1 | Definitive | 81.03 | NM_014991.6 | Very Strong | PMID: 31327001:Wdfy3-haploinsufficient mice show macrocephaly, motor deficits, and learning impairments, supporting loss-of-function as a disease mechanism and application of PVS1. |
| chr4-113354556-A-T | ANK2 | stop gained | PVS1 | Definitive | 87.5 | NM_001148.6 | Very Strong | PMID: 37195288:ANK2 haploinsufficiency is an established disease mechanism, supported by de novo truncating variants in NDD cases and functional studies showing neuronal hyperexcitability, justifying application of PVS1. |
| chr4-113359220-C-CTGTGTCAGCCT | ANK2 | frameshift variant | PVS1 | Definitive | 87.5 | NM_001148.6 | Very Strong | PMID: 37195288:ANK2 haploinsufficiency is an established disease mechanism, supported by de novo truncating variants in NDD cases and functional studies showing neuronal hyperexcitability, justifying application of PVS1. |

| | | | | | | | | |
|---|---|---|---|---|---|---|---|---|
| chr4-113369636-CCCCAACA-C | ANK2 | frameshift variant | PVS1 | Definitive | 87.5 | NM_001148.6 | Very Strong | PMID: 37195288:ANK2 haploinsufficiency is an established disease mechanism, supported by de novo truncating variants in NDD cases and functional studies showing neuronal hyperexcitability, justifying application of PVS1. |
| chr4-113369647-AGCGAGCGGGGAGG-A | ANK2 | frameshift variant | PVS1 | Definitive | 87.5 | NM_001148.6 | Very Strong | PMID: 37195288:ANK2 haploinsufficiency is an established disease mechanism, supported by de novo truncating variants in NDD cases and functional studies showing neuronal hyperexcitability, justifying application of PVS1. |
| chr4-113369662-TCTCCC-T | ANK2 | frameshift variant | PVS1 | Definitive | 87.5 | NM_001148.6 | Very Strong | PMID: 37195288:ANK2 haploinsufficiency is an established disease mechanism, supported by de novo truncating variants in NDD cases and functional studies showing neuronal hyperexcitability, justifying application of PVS1. |
| chr4-113369669-TCA-T | ANK2 | frameshift variant | PVS1 | Definitive | 87.5 | NM_001148.6 | Very Strong | PMID: 37195288:ANK2 haploinsufficiency is an established disease mechanism, supported by de novo truncating variants in NDD cases and functional studies showing neuronal hyperexcitability, justifying application of PVS1. |
| chr4-113369677-GAACCCGAAGA-G | ANK2 | frameshift variant | PVS1 | Definitive | 87.5 | NM_001148.6 | Very Strong | PMID: 37195288:ANK2 haploinsufficiency is an established disease mechanism, supported by de novo truncating variants in NDD cases and functional studies showing neuronal hyperexcitability, justifying application of PVS1. |
| chr4-113369689-C-CTGGTTTT | ANK2 | frameshift variant | PVS1 | Definitive | 87.5 | NM_001148.6 | Very Strong | PMID: 37195288:ANK2 haploinsufficiency is an established disease mechanism, supported by de novo truncating variants in NDD cases and functional studies showing neuronal hyperexcitability, justifying application of PVS1. |
| chr4-113369691-CTCAGA-C | ANK2 | frameshift variant | PVS1 | Definitive | 87.5 | NM_001148.6 | Very Strong | PMID: 37195288:ANK2 haploinsufficiency is an established disease mechanism, supported by de novo truncating variants in NDD cases and functional studies showing neuronal hyperexcitability, justifying application of PVS1. |
| chr4-113369698-CACAGAGA-C | ANK2 | frameshift variant | PVS1 | Definitive | 87.5 | NM_001148.6 | Very Strong | PMID: 37195288:ANK2 haploinsufficiency is an established disease mechanism, supported by de novo truncating variants in NDD cases and functional studies showing neuronal hyperexcitability, justifying application of PVS1. |
| chr5-14487813-CG-C | TRIO | frameshift variant | PVS1 | Definitive | 76.71 | NM_007118.4 | Very Strong | PMID: 30840899:TRIO haploinsufficiency causes behavioral, structural, and synaptic deficits in mice relevant to ASD, supporting loss-of-function as a disease mechanism and application of PVS1. |
| chr6-156778291-A-AGC | ARID1B | frameshift variant | PVS1 | Definitive | 94.05 | NM_001374828.1 | Very Strong | PMID: 29184203:ARID1B haploinsufficiency is a validated disease mechanism, with functional studies in mouse models demonstrating neurodevelopmental and behavioral deficits relevant to ASD, supporting application of PVS1. |
| chr6-156778292-A-AGCAG | ARID1B | frameshift variant | PVS1 | Definitive | 94.05 | NM_001374828.1 | Very Strong | PMID: 29184203:ARID1B haploinsufficiency is a validated disease mechanism, with functional studies in mouse models demonstrating neurodevelopmental and behavioral deficits relevant to ASD, supporting application of PVS1. |
| chr6-156778292-A-AG | ARID1B | frameshift variant | PVS1 | Definitive | 94.05 | NM_001374828.1 | Very Strong | PMID: 29184203:ARID1B haploinsufficiency is a validated disease mechanism, with functional studies in mouse models demonstrating neurodevelopmental and behavioral deficits relevant to ASD, supporting application of PVS1. |
| chr6-156778292-A-AGCAGCAG | ARID1B | frameshift variant | PVS1 | Definitive | 94.05 | NM_001374828.1 | Very Strong | PMID: 29184203:ARID1B haploinsufficiency is a validated disease mechanism, with functional studies in mouse models demonstrating neurodevelopmental and behavioral deficits relevant to ASD, supporting application of PVS1. |
| chr7-152162427-T-TATGACTGTTTTCTTTCTACACAG | KMT2C | stop gained & frameshift variant | PVS1 | Definitive | 86.99 | NM_170606.3 | Very Strong | PMID: 37538398:Brain-specific KMT2C knockout mice show ASD-like behaviors and cognitive deficits, supporting loss-of-function as a disease mechanism and application of PVS1. |

| | | | | | | | | |
|---|---|---|---|---|---|---|---|---|
| chr7-152177898-A-AGCTTGCCAGT | KMT2C | frameshift variant | PVS1 | Definitive | 86.99 | NM_170606.3 | Very Strong | PMID: 37538398:Brain-specific KMT2C knockout mice show ASD-like behaviors and cognitive deficits, supporting loss-of-function as a disease mechanism and application of PVS1. |
| chr7-152187302-G-T | KMT2C | stop gained | PVS1 | Definitive | 86.99 | NM_170606.3 | Very Strong | PMID: 37538398:Brain-specific KMT2C knockout mice show ASD-like behaviors and cognitive deficits, supporting loss-of-function as a disease mechanism and application of PVS1. |
| chr7-152194508-G-T | KMT2C | stop gained | PVS1 | Definitive | 86.99 | NM_170606.3 | Very Strong | PMID: 37538398:Brain-specific KMT2C knockout mice show ASD-like behaviors and cognitive deficits, supporting loss-of-function as a disease mechanism and application of PVS1. |
| chr9-106926840-CCTGA-C | ZNF462 | frameshift variant | PVS1 | Definitive | 97.14 | NM_021224.6 | Very Strong | PMID: 27621227:ZNF462 haploinsufficiency is supported by knockout mouse models showing neurodevelopmental and behavioral abnormalities, validating loss-of-function as a disease mechanism and supporting application of PVS1. |
| chr9-106926847-G-T | ZNF462 | stop gained | PVS1 | Definitive | 97.14 | NM_021224.6 | Very Strong | PMID: 27621227:ZNF462 haploinsufficiency is supported by knockout mouse models showing neurodevelopmental and behavioral abnormalities, validating loss-of-function as a disease mechanism and supporting application of PVS1. |
| chr9-106927204-T-TC | ZNF462 | frameshift variant | PVS1 | Definitive | 97.14 | NM_021224.6 | Very Strong | PMID: 27621227:ZNF462 haploinsufficiency is supported by knockout mouse models showing neurodevelopmental and behavioral abnormalities, validating loss-of-function as a disease mechanism and supporting application of PVS1. |
| chr9-106927204-T-TGC | ZNF462 | frameshift variant | PVS1 | Definitive | 97.14 | NM_021224.6 | Very Strong | PMID: 27621227:ZNF462 haploinsufficiency is supported by knockout mouse models showing neurodevelopmental and behavioral abnormalities, validating loss-of-function as a disease mechanism and supporting application of PVS1. |
| chr9-106927204-T-TGCCCCCCCCCC | ZNF462 | frameshift variant | PVS1 | Definitive | 97.14 | NM_021224.6 | Very Strong | PMID: 27621227:ZNF462 haploinsufficiency is supported by knockout mouse models showing neurodevelopmental and behavioral abnormalities, validating loss-of-function as a disease mechanism and supporting application of PVS1. |
| chr9-106927221-A-ACCCCCCCC | ZNF462 | frameshift variant | PVS1 | Definitive | 97.14 | NM_021224.6 | Very Strong | PMID: 27621227:ZNF462 haploinsufficiency is supported by knockout mouse models showing neurodevelopmental and behavioral abnormalities, validating loss-of-function as a disease mechanism and supporting application of PVS1. |
| chr9-137744014-A-AGCAGGCGCGCGCGCGGCGAGACCTTCTTGGTCAGGTCCTCGATCAGTCCCGAGGCGAAGGCCGGGATGCC | EHMT1 | frameshift variant | PVS1 | Definitive | 83.19 | NM_024757.5 | Very Strong | PMID: 28742076:Patient-derived hiPSC neurons with EHMT1 loss-of-function show reduced expression, impaired neurite morphology, and altered neuronal activity, supporting haploinsufficiency as a disease mechanism in Kleefstra syndrome associated with ASD and supporting application of PVS1. |
| chrX-41216447-C-CTGCT | USP9X | frameshift variant | PVS1 | Definitive | 68.14 | NM_001039591.3 | Very Strong | PMID: 23861879: Loss of USP9X in neural tissue leads to structural brain abnormalities and ASD-like behaviors in forebrain-specific knockout mice, supporting LoF as a pathogenic mechanism in females and justifying application of PVS1. |
| chrX-74740391-G-GATATATGATTAACAAATATTTTCTACCATTCCGTGGTTTGTGTTTTCATT | NEXMIF | stop gained & frameshift variant | PVS1 | Definitive | 99.5 | NM_001008537.3 | Strong | PMID: 31704787:The NEXMIF KO mice demonstrate autism-like behaviors including deficits in social interaction, increased repetitive self-grooming, and impairments in communication and in learning and memory. |
| chrX-147928713-A-ATT | FMR1 | frameshift variant | PVS1 | Definitive | 59.09 | NM_002024.6 | Very Strong | PMID:8033209, PMID: 34440392:Fmr1 knockout mice lack FMRP and exhibit key phenotypes such as learning deficits, macroorchidism, and hyperactivity, supporting FMR1 loss-of-function as a disease mechanism relevant to ASD and justifying application of PVS1. |
| chrX-147928716-G-T | FMR1 | stop gained | PVS1 | Definitive | 59.09 | NM_002024.6 | Very Strong | PMID:8033209, PMID: 34440392:Fmr1 knockout mice lack FMRP and exhibit key phenotypes such as learning deficits, macroorchidism, and hyperactivity, supporting FMR1 loss-of-function as a disease mechanism relevant to ASD and justifying application of PVS1. |
